# Investigating a causal role for neutrophil count on *P. falciparum* severe malaria: a Mendelian Randomization study

**DOI:** 10.1101/2023.09.06.23295065

**Authors:** Andrei-Emil Constantinescu, David A. Hughes, Caroline J. Bull, Kathryn Fleming, Ruth E. Mitchell, Jie Zheng, Siddhartha Kar, Nicholas J. Timpson, Borko Amulic, Emma E. Vincent

**Affiliations:** MRC Integrative Epidemiology Unit at the University of Bristol, Bristol, UK; Bristol Medical School, Population Health Sciences, University of Bristol, Bristol, UK; School of Translational Health Sciences, University of Bristol, Bristol, UK; School of Cellular and Molecular Medicine, University of Bristol, Bristol, UK; Department of Endocrine and Metabolic Diseases, Shanghai Institute of Endocrine and Metabolic Diseases, Ruijin Hospital, Shanghai Jiao Tong University School of Medicine, Shanghai, China; Shanghai National Clinical Research Center for Metabolic Diseases, Key Laboratory for Endocrine and Metabolic Diseases of the National Health Commission of the PR China, Shanghai National Center for Translational Medicine, Ruijin Hospital, Shanghai Jiao Tong University School of Medicine, Shanghai, China

**Author notes:** Corresponding authors Correspondence to Borko Amulic and Emma E. Vincent. Joint senior authors (Borko Amulic, Emma E. Vincent).

**Keywords:** Malaria, Neutrophil count, Mendelian randomization, GWAS, African ancestry

## Abstract

**Background:** Malaria caused by *P. falciparum* imposes a tremendous public health burden on people living in sub-Saharan Africa. Severe malaria is associated with high morbidity and mortality and results from complications such as cerebral malaria, severe anaemia or respiratory distress. Individuals living in malaria endemic regions often have a reduced circulating neutrophil count due to a heritable phenomenon called ‘benign ethnic neutropenia’ (BEN). Neutrophils defend against bacterial infections but have been shown to be detrimental in pre-clinical malaria models, raising the possibility that reduced neutrophil counts modulate severity of malaria in susceptible populations. We tested this hypothesis by performing a genome-wide association study (GWAS) of circulating neutrophil count and a Mendelian randomization (MR) analysis of neutrophil counts on severe malaria in individuals of predominantly African ancestry.

**Results:** We carried out a GWAS of neutrophil count in individuals associated to an African continental ancestry group within UK Biobank (N=5,976). We identified previously unknown loci regulating neutrophil count in a non-European population. This was followed by a two-sample bi-directional MR analysis between neutrophil count and severe malaria (MalariaGEN, N=17,056). We identified 73 loci (r^2^=0.1) associated with neutrophil count, including the well-known rs2814778 variant responsible for BEN. The greatest evidence for an effect was found between neutrophil count and severe anaemia, although the confidence intervals crossed the null. MR analyses failed to suggest evidence for an effect of the combined severe malaria syndromes or individual subtypes (severe malaria anaemia, cerebral malaria, other severe malaria) on neutrophil count.

**Conclusion:** Our GWAS of neutrophil count revealed unique loci present in individuals of African ancestry. We note that a small sample-size reduced our power to identify variants with low allele frequencies and/or low effect sizes in our GWAS. Our work highlights the need for conducting large-scale biobank studies in Africa and for further exploring the link between neutrophils and severe malarial anemia.

## Introduction

Malaria is a mosquito-transmitted disease that annually affects approximately 215 million people [1,2]. The disease is caused by protozoan parasites of the *Plasmodium* genus: *Plasmodium falciparum* (*P. falciparum*) causes life-threatening disease in sub-Saharan Africa and accounts for almost all malaria deaths, while *P. vivax* leads to a milder disease that is nonetheless associated with a significant public health burden in diverse geographical regions [2].

*P. falciparum* malaria causes approximately 400,000-600,000 deaths each year, primarily in African children under the age of five [1]. The majority of *P. falciparum* malaria cases consist of uncomplicated febrile illness, however a portion of nonimmune infected individuals succumb to severe malaria, which can manifest as cerebral malaria, severe anemia, acute respiratory distress or kidney injury [3]. *Plasmodium* resides and proliferates in red blood cells (RBCs) and pathology is triggered by cytoadherence of infected RBCs (iRBCs) to microcapillary endothelia in different organs, which can lead to vascular occlusion and endothelial permeability [3]. Inflammation plays a key role in both facilitating iRBC sequestration [4] and in tissue damage [3,5,6]. In cerebral malaria, the deadliest form of the disease, iRBCs sequester in the neurovasculature, provoking blood brain barrier permeabilization, vascular leak and brain swelling [3].

Malaria has been the biggest cause of childhood deaths over the past 5000 years [7]. As such, it has exerted the strongest known selective pressure on the human genome and has resulted in the selection of various polymorphisms that confer *Plasmodium* tolerance or resistance. Among the most prominent examples are haemoglobin S (Hbs; sickle cell trait) [8] and alpha-thalassemia variants [9], both of which are common in malaria endemic regions despite causing disease in the homozygous state [7]. The HbS polymorphism in the heterozygous state confers the greatest protection (effect size >80%; [7,10]). The heritability of severe malaria is estimated to be around 30% [11,12] but the cumulative effect of the aforementioned variants is thought to only be 2% [7,11], suggesting that polygenic interactions may account for a large part of the missing heritability of this complex disease.

Individuals living in malaria-endemic regions, as well as those descended from them, often have reduced numbers of neutrophils in circulation as compared to those living in non- endemic regions. This heritable phenomenon is called ‘benign ethnic neutropenia’ (BEN) and is distinct from life-threatening severe neutropenia. BEN is prominent in South Mediterranean, Middle Eastern, sub-Saharan African and West Indies populations [13]. BEN is estimated to occur in 25-50% of Africans [13–15] and 10.7% of Arabs [16] but in less than 1% of people of European ancestry living in the Americas [17]. Neutrophils are essential for immune defense against bacteria and fungi [18], however BEN does not lead to significantly greater susceptibility to infection in the United States [13]. Nevertheless, it remains curious that selection for lower neutrophil counts occurred in sub-Saharan Africa, a region associated with a high infectious disease burden. This observation is partly explained by the finding that in populations of African and Yemenite Jewish ancestry, BEN is strongly associated with a polymorphism in the atypical chemokine receptor 1 (ACKR1/DARC), which encodes the Fy/Duffy antigen, a surface receptor utilized by *P. vivax* to invade RBCs [19]. This variant abolishes expression of ACKR1 on RBCs and is thought to contribute to low prevalence of *P. vivax* in sub-Saharan Africa, where the polymorphism is found at levels close to fixation [7]. ACKR1, in addition to serving as one of the invasion receptors for *P. vivax*, controls circulating levels of chemokines [20], which also regulate blood neutrophil numbers [20]. It is unclear to what extent other polymorphisms contribute to BEN in individuals living in malaria endemic regions [21].

Neutrophils have recently been shown to have a detrimental role in malaria, promoting pathogenesis by enhancing sequestration of iRBCs [4] and contributing to inflammatory tissue damage [6,22,23]. Altered neutrophil responses have also been linked to severe malarial anemia in paediatric patients [24]. On the other hand, neutrophils have also been suggested to participate in parasite clearance [25] and in shaping the *Plasmodium* antigenic repertoire [26]. These studies raise the possibility that neutropenia in malaria endemic regions may modulate severity of *P. falciparum* malaria, in addition to conferring resistance to *P. vivax*. However, observational studies, such as the ones referenced above, are prone to confounding and reverse causation [27–29]. It is therefore essential to employ additional methods, such as those in population genetics, to study the link between neutrophil count and *P. falciparum* severe malaria, with the overarching aim to improve the health outcomes of the people residing in endemic regions.

Mendelian randomization (MR) is a method in genetic epidemiology which uses genetic variants as proxies with the aim of providing evidence for causal inference between an exposure and an outcome [27]. As the majority of alleles are assigned randomly at birth, an MR analysis is analogous to that of a randomized control trial (RCT), the most reliable method for evaluating the effectiveness of an intervention [30]. Large-scale studies, such as UK Biobank (UKBB) [31], have increased the potential of MR studies due to the increase in power to detect associations in genome-wide association studies (GWASs) that comes with such a large sample size.

Recent efforts in genetics have resulted in the generation of hundreds of GWAS using UKBB’s non-European participants for many traits in a hypothesis-free manner (https://pan.ukbb.broadinstitute.org/). However, the same covariates were used for each trait, and the impact of confounding due to population structure was not studied, this represents a potential limitation for constructing reliable instruments for a MR analysis [32]. A recent study by Chen et al. used individuals of non-European ancestry in UKBB to perform trans-ancestry GWAS of blood cell traits (BCTs) [33]. However, the African continental ancestry groups (CAGs) of UKBB display strong population structure [34]. It therefore remains unclear whether a GWAS of a complex trait, such as neutrophil count, would result in associations that are linked to a biological mechanism, or whether the associations would be a product of confounding due to residual population structure. In order to answer these questions, a more thorough investigation of the sampled dataset is warranted. This becomes even more important when aiming to conduct causal inference analyses in genetic epidemiology, such as two-sample Mendelian randomization [35,36].

To test the hypothesis that reduced neutrophil counts modulates severity of malaria in susceptible populations, we first performed a GWAS of neutrophil count in individuals associated to the UKBB African continental ancestry group (CAG), described in our previous study [34]. Here, we conducted a series of sensitivity analyses to describe the GWAS results and selection of genetic instruments to proxy for neutrophil count in a MR analysis. We then conducted bi-directional MR to estimate the casual relationship between neutrophil count and SM using data from the MalariaGEN consortium [37].

## Materials & Methods

### Study design

6,653 people representing the UKBB African CAG were identified as part of our previous study [34]. After PCA outlier filtering [34], we also excluded those without neutrophil count data and blood-related disorders [38], resulting in a final sample of 5,976. The primary GWAS of neutrophil count used in all other analyses was generated with BOLT-LMM. Several analyses were undertaken afterwards to test the validity of the primary GWAS estimates. Following this, an MR analysis was performed between neutrophil count and severe malaria caused by *P. falciparum* using data from MalariaGEN (**Figure 1**).

**Figure 1.**
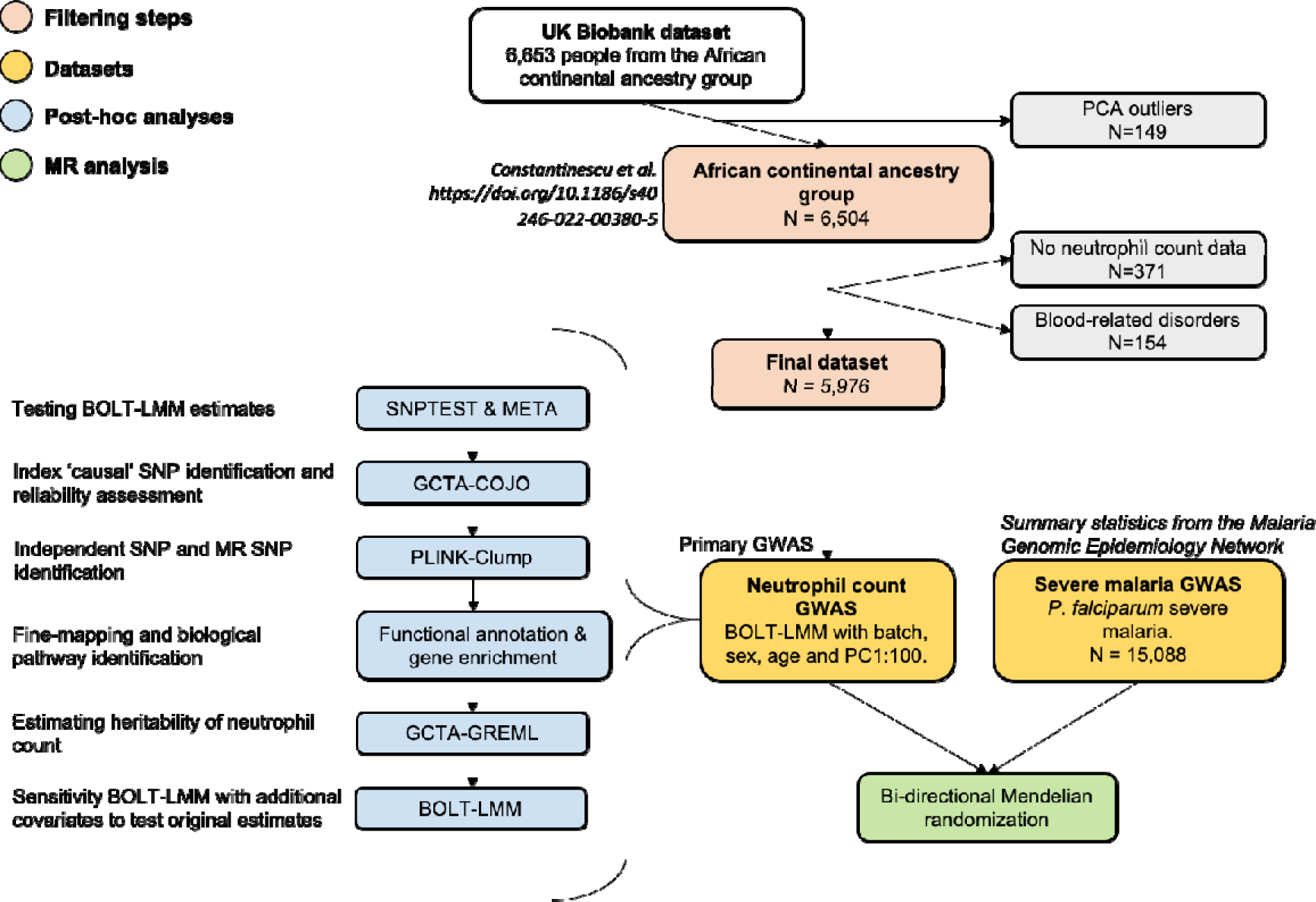
Study design of the project.

### UK Biobank genetic data

UK Biobank’s “non-white” British data was studied previously, where 6,653 people corresponded to the African CAG, of which 6,504 remained (5,989 unrelated; 515 related) after filtering for principal component analysis (PCA) outliers [34]. These were further assigned into seven clusters based on a K-means clustering algorithm (K1=527; K2=1,177; K3=1,176; K4=1,001; K5=1,206; K6=862; K7=184) [34]. This dataset (N=6,504) included both directly genotyped (N=784,256) and imputed (N=29,363,284) SNPs filtered with a minor allele count of >20. We filtered out SNPs with an INFO score threshold of 0.3, as it gives the best balance between data quality and quantity. Another filtering process was a Hardy-Weinberg equilibrium (HWE) test (P<1e-10), used to identify SNPs of poor genotyping quality [39]. Finally, related individuals from the dataset were removed, resulting in 5,509 unrelated people in the filtered African CAG dataset. SNPs with a minor allele count of less than 17 (corresponding to the new sample-size from 20) were removed. 23,530,028 SNPs remained after filtering by INFO score, HWE test and minor allele count.

### UK Biobank phenotypic data

Haematological samples were analysed using four Beckman Coulter LH750 instruments [40]. Total white blood cell (WBC) count and neutrophil percentage (%) were measured through the Coulter method, with neutrophil count derived as “neutrophil % / 100 x total WBC” and expressed as 10^9^ cells/Litre [40]. Afterwards, the sample collection date was split into year, month, day, and minutes (passed since the start of the day of the appointment visit), while the neutrophil count measurement variable was log-transformed into a variable named “nc_log”, which was used as the default neutrophil count variable throughout the study. Other variables that were used in the main analyses were: age, genetic sex, blood sample device ID, UKBB assessment centre and principal components (PCs) 1 to 100. Filtering was performed based on the selection criteria described by Astle et al. [38] and Chen et al. [33]. Briefly, individuals with disorders/diseases that could affect blood counts (e.g. HIV, leukaemia, congenital anaemias, cirrhosis) were removed, bringing the final sample size to 5,976. This dataset is referred to as “AFR_CAG”.

### BOLT-LMM GWAS

BOLT-LMM was used as the software to run the primary (main) GWAS. Linkage disequilibrium (LD) scores were generated from the directly genotyped dataset that is required by BOLT-LMM to calibrate the test statistics. After preparing the phenotypic data to match the desired input, BOLT-LMM was run on AFR_CAG adjusting for age, genetic sex, UKBB assessment centre, blood sampling device, sampling year, sampling month, sampling day, minutes passed in sampling day and the first 100 principal components (PCs). Two linear model GWAS in SNPTEST were also completed on each K-means cluster and then meta-analysed: one without accounting for the Duffy SNP rs2814778 called “META-WOD”, and one where the Duffy SNP was included as a covariate, called “META-WD”. Another BOLT-LMM sensitivity run was done with additional covariates to further study the validity of the main GWAS findings (**Supplementary Methods**).

### Conditional & joint association analysis

We used GCTA-COJO [41,42] to identify independent signals from the BOLT-LMM GWAS, as well as to detect any possible secondary signals arising from a stepwise selection model. SNPs which are close together are usually in LD i.e. their alleles are not random, but correlated [39]. Before running GCTA-COJO, related individuals were filtered out of the dataset. PLINK was then used on this resulting output to perform a greedy filtering of related individuals. Following this step, GCTA-COJO was run on the AFR_CAG filtered dataset to identify conditionally independent SNPs. These were referred to as “index” SNPs in the text.

### PLINK clumping

After GCTA-COJO, we used PLINK to perform clumping with three different thresholds. The first two represent the thresholds for defining LD independent SNPs for running analyses on the online variant annotation platform Functional Mapping and Annotation (FUMA) [43], while the latter being the clumping conditions used for conducting a Mendelian randomization analysis [44,45].

1. --clump-p1=5e-8, --clump-r2=0.6, --clump-kb=250
2. --clump-p1=5e-8, --clump-r2=0.1, --clump-kb=250
3. --clump-p1=5e-8, --clump-r2=0.001, --clump-kb=10000

### Heritability analysis

An analysis was conducted with GCTA-GREML to estimate the proportion of variance in neutrophil count explained by all genetic variants present in the filtered AFR_CAG dataset [46], with and without adjusting for the Duffy SNP rs2814778.

### *P. falciparum* severe malaria genetic data

GWAS summary statistics for *P. falciparum* severe malaria were downloaded from a case-control study that spanned nine African and two Asian countries [37]. In brief, controls samples were gathered from cord blood, and in some cases, from the general population. Cases were diagnosed according to WHO definitions of severe malaria [47] and were categorised according to CM, severe malarial anemia (SMA) and other severe malaria (OTHER) symptoms (**Supplementary Table 1**). The majority of the RSIDs in the MalariaGEN dataset used older identifiers, and some of them had the “kgp” prefix that comes with the Illumina-HumanOmni2.5M array. Ideally, in a two-sample MR setting, the two samples would have a perfect match in the available genetic variants. It is desirable to at least maximise the number of matching variants to test. Therefore, RSID information for the MalariaGEN variants was updated in R by using the filtered AFR_CAG dataset as a reference panel.

### Meta-analysis of severe malaria African populations

Summary statistics for severe malaria and its sub-phenotypes were generated from a meta-analysis which included individuals from two non-African countries – Vietnam and Papua New Guinea. The inclusion of SNP effect sizes from GWAS conducted in heterogenous population might bias MR estimates [48]. Therefore, per-population summary statistics were downloaded (https://www.malariagen.net/sppl25/) for each African country in the study and a meta-analysis was conducted on them using METAL [49–51].

### Mendelian randomization analysis

The “TwoSampleMR” R package [52,53] was used to perform the MR analyses. The two datasets were harmonised i.e. orientated on the same strand and if SNPs were not found in the outcome dataset, we searched for SNP proxies. We then conducted a bi-directional MR analysis, where the effect of neutrophil count on overall severe malaria, along with the three sub-phenotypes was estimated and vice-versa. The main analysis was conducted using an IVW model [54]. Additionally, we ran a sensitivity MR analysis to outline the effect estimates of each SNP on the desired outcome, with IVW and MR-Egger [55,56] estimates where the number of instruments was larger than two and three, respectively.

## Results

### Analysis of study sample

5,976 out of 6,504 individuals in AFR_CAG remained after filtering for missing data and traits affecting blood cells. The mean value for neutrophil count was 2.9 x 10^9^ cells/litre, as expected this was lower than a European sample (4.21 x 10^9^ cells/Litre) [33,38]. The GWAS sample had a larger proportion of females (57%), was of a higher mean age (39 vs. 58.1 years) [57] and slightly higher body mass index (BMI) (27.6 vs. 29.8 kg/m^2^) [58] than the general UK population (**Table 1**).

**Table 1.**
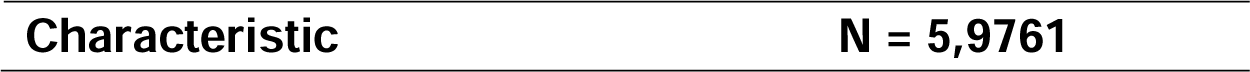
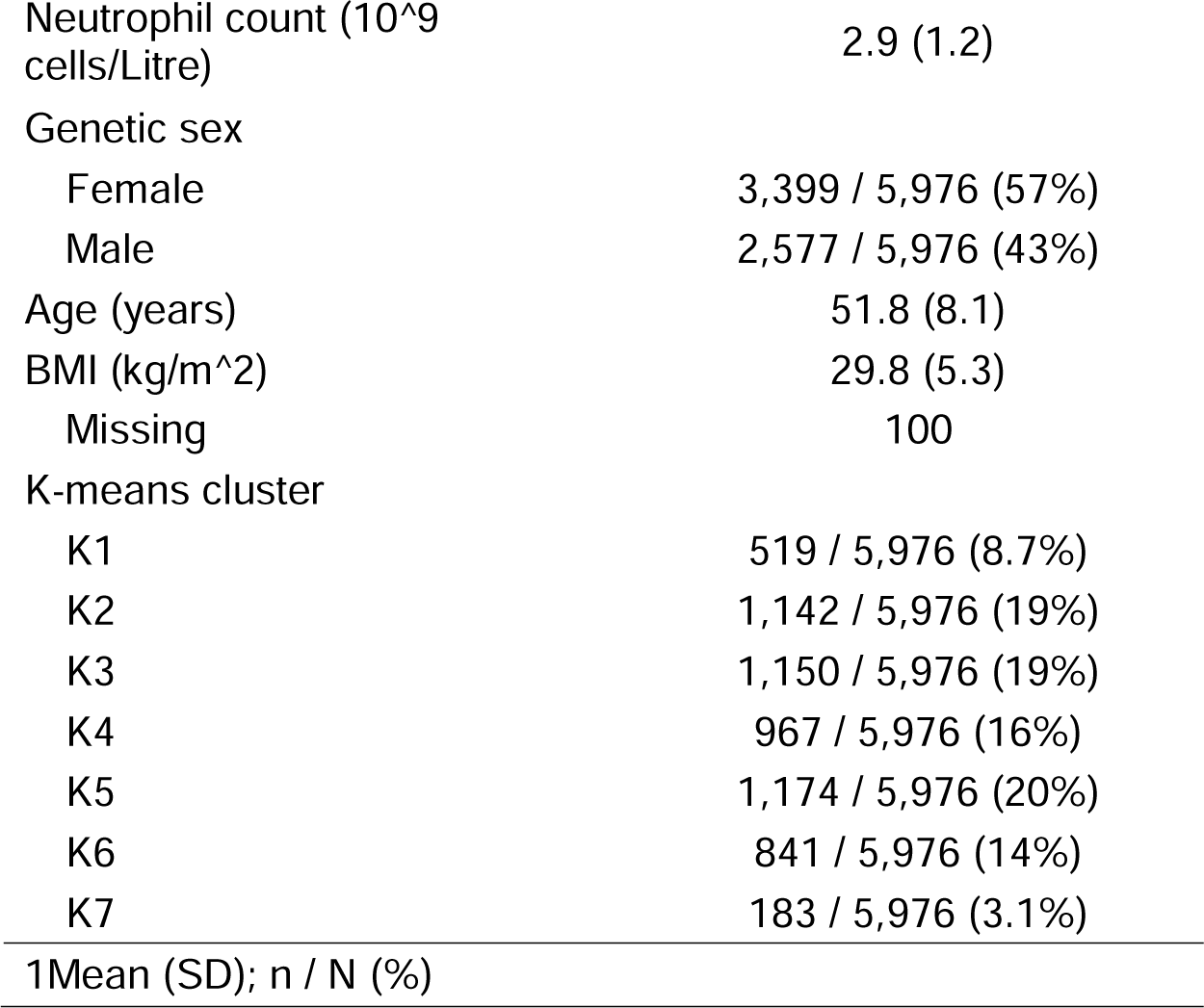
Description of GWAS sample.

| Characteristic | N = 5,9761 |
| --- | --- |
| Neutrophil count (10<sup>9</sup> cells/Litre) | 2.9 (1.2) |
| Genetic sex |  |
| Female | 3,399 / 5,976 (57%) |
| Male | 2,577 / 5,976 (43%) |
| Age (years) | 51.8 (8.1) |
| BMI (kg/m<sup>2</sup>) | 29.8 (5.3) |
| Missing | 100 |
| K-means cluster |  |
| K1 | 519 / 5,976 (8.7%) |
| K2 | 1,142 / 5,976 (19%) |
| K3 | 1,150 / 5,976 (19%) |
| K4 | 967 / 5,976 (16%) |
| K5 | 1,174 / 5,976 (20%) |
| K6 | 841 / 5,976 (14%) |
| K7 | 183 / 5,976 (3.1%) |
| 1Mean (SD); n / N (%) |  |

We used the natural log-transformation, nc_log, in the GWAS. There was some variation in nc_log between each K-means cluster (Kpop) (**Figure 2B**), although this was low, with the median hovering around 1 (**Figure 2A**).

**Figure 2.**
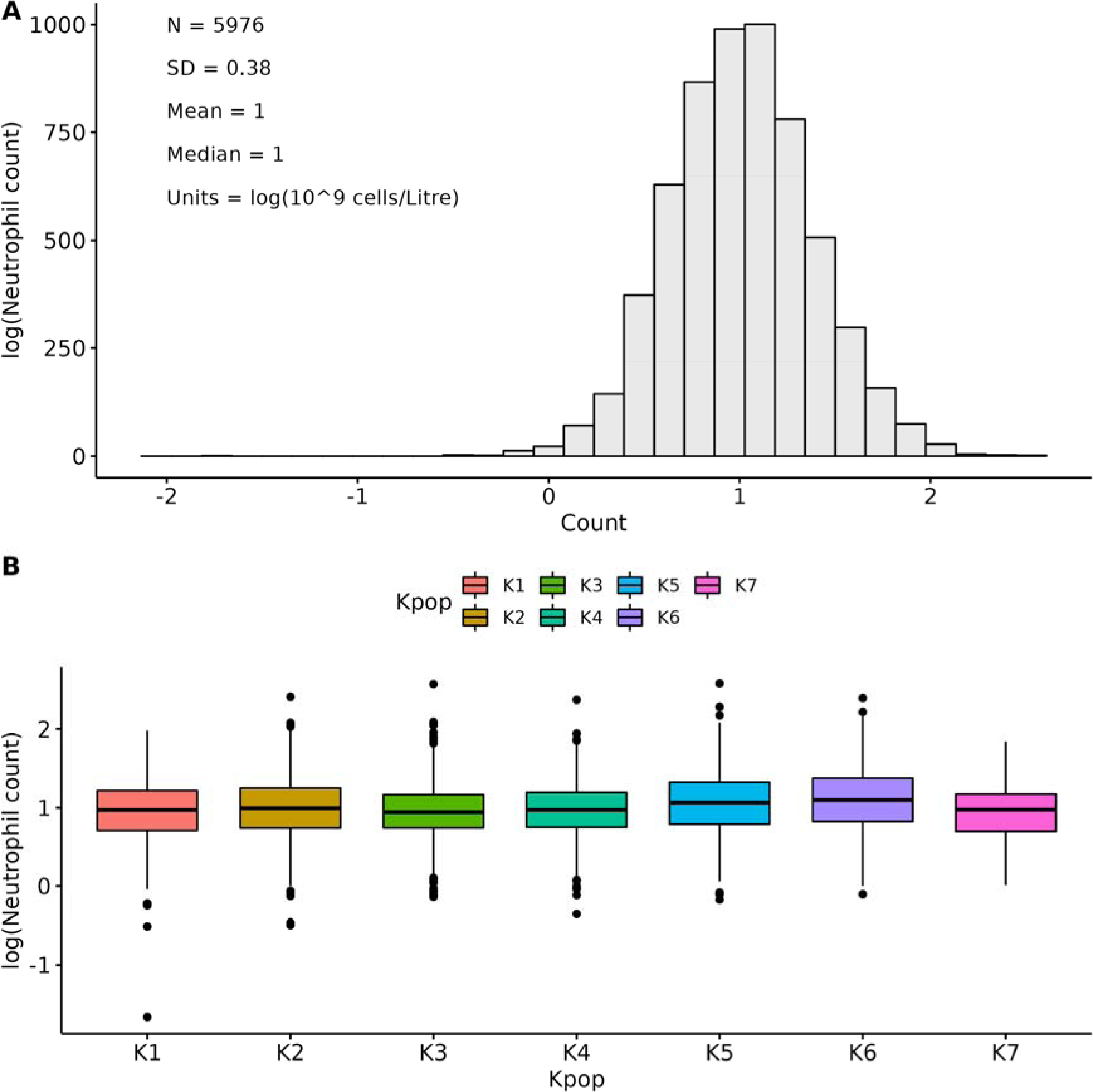
Neutrophil count variation in the GWAS sample. A histogram outlining the distribution of neutrophil levels is shown in the whole AFR_CAG population (**A**), along with representative boxplots describing neutrophil count variation by K-means cluster sample (**B**).

GWAS are usually performed using individuals of a similar genetic background to avoid SNP-trait associations that are biased or are false-positives due to a confounding effect by ancestry [59]. However, even in white British individuals from UKBB, latent population structure can affect SNP effect sizes, which may not be completely removed by adjusting for PCs [60]. Nevertheless, we investigated the number of PCs that should be added into the GWAS to control for population structure. The Tracy-Widom statistic from the EIGENSOFT package [61] indicated over 100 significant PCs. However, there is no exact way to establish how many PCs should be added into a GWAS, although an excessive number of PCs can lead to a reduction in power, while too few might bias GWAS effect sizes due to residual population structure [48]. Previous studies have added 40 to 100 PCs in their UKBB GWA analyses [60,62,63], hence our inclusion of the first 100 PCs as covariates.

Next, we conducted a power calculation supposing a linear, additive, GWA model. Sample- size is an important factor in GWAS [64,65], and the higher the sample-size, the higher the power to detect SNPs which explain a smaller proportion of the variance (heritability) in a particular trait [66]. The power to detect an association was >80% when the proportion of variance explained by SNPs was higher than 0.75% (**Supplementary Figure 1**).

### Genome-wide association study

We used BOLT-LMM for the main GWAS, which employs a linear-mixed model algorithm for conducting association testing [67]. It is unknown how well linear mixed model using PCs and kinship matrixes performs in highly stratified population samples with complex demographic histories and unique allele frequencies and linkage disequilibrium [68]. To ensure that results derived by a linear mixed model as implemented by BOLT-LMM were reliable, we also aimed to conduct additional GWAS using a standard linear model on less stratified sub-samples of our sample population – as identified using an unsupervised machine learning methodology (**Supplementary Methods**).

This AFR_CAG filtered sample was taken forward for further analyses. 704 genetic variants passed the GWAS significance threshold of P < 5e-8 in the primary GWAS. Most of these signals were in chromosome 1, in the proximity of the ACKR1-associated rs2814778, which had the lowest P-value across the genome (2.7E-87) (**Figure 3A**). The META-WOD GWAS had 373 variants passing the threshold, while the META-WD (with Duffy adjustment) GWAS had 31 significant SNPs, evidencing that most of the identified top signals in META- WOD were likely in LD with rs2814778. The QQ-plot of the BOLT-LMM GWAS did not display an early deviation from the expected P-value, indicating low likelihood of systemic bias in association statistics [69] (**Figure 3B**).

**Figure 3.**
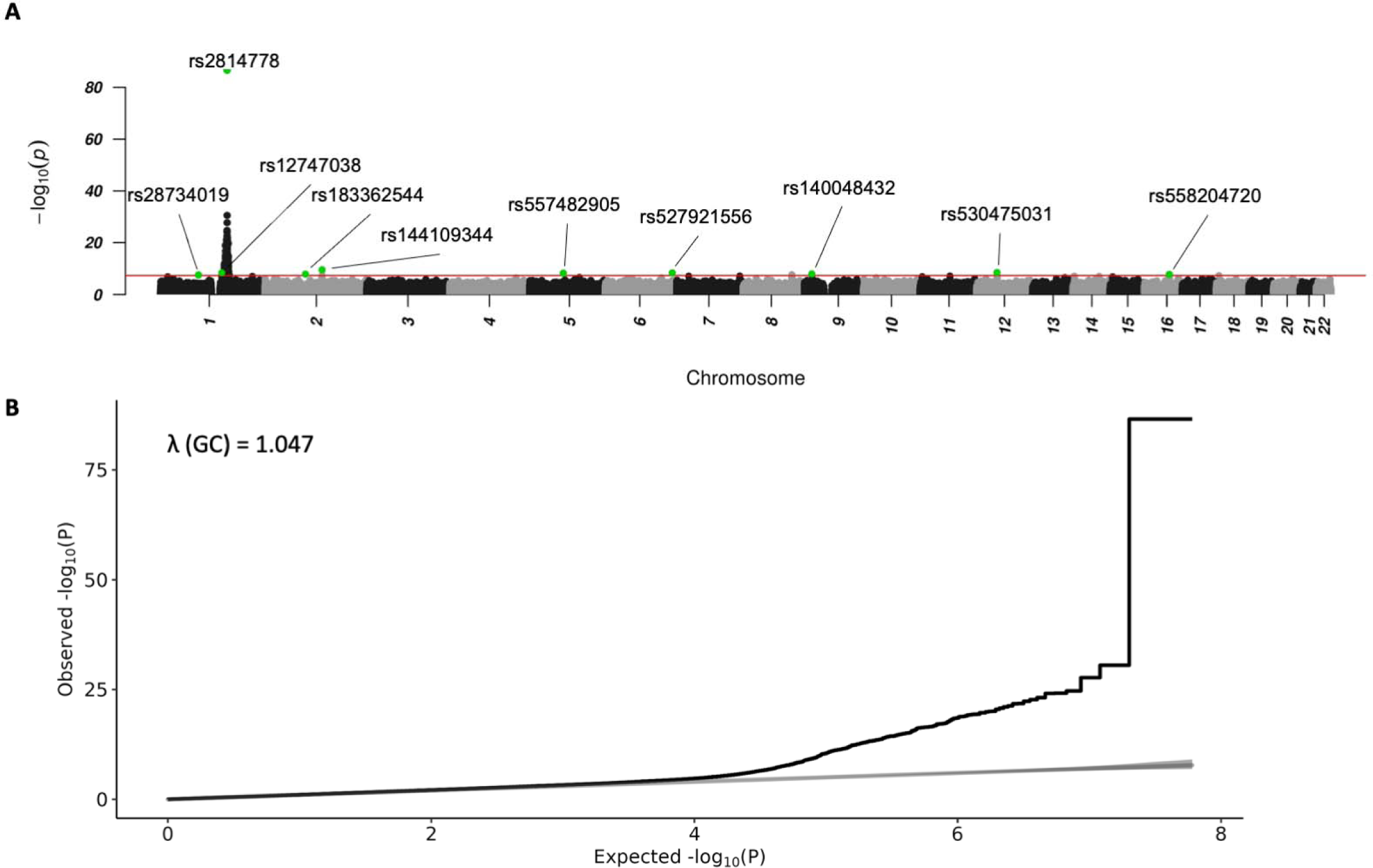
Manhattan plot of neutrophil count GWAS. The x-axis is the base-pair position inside each chromosome, while the y-axis is the -log of the association P-value. A GWAS significance line is drawn to correspond to P=5e-8 on the -log(P) axis (**A**). Index SNPs from the GCTA-COJO run are highlighted in green. QQ-Plot of observed vs. expected P-values for each SNP, along with the genomic inflation factor on the top-left (B).

Next, we aimed to identify which SNPs might causally associate with neutrophil count. To do this, we used a conservative GCTA-COJO approach [42], which yielded 10 index SNPs (**Figure 3**, **Table 2**). Genomic location context of each index SNP is available in Supplementary Figures 2-4.

**Table 2.** GCTA-COJO index SNPs. BETA, SE and P BOLT are the regression statistics of the BOLT-LMM neutrophil count GWAS. BETA, SE and P for META-WOD and META-WD are the regression statistics of the meta-analysed GWAS done on each Kpop, without and with adjustment for the Duffy SNP, respectively, META-N and META-N-Studies indicate the number of meta-analysed individuals in each Kpop, and the number of Kpops included in the meta-analysis.

| SNP | BETA-BOLT | SE-BOLT | P-BOLT | BETA-META-WOD | BETA-META-WD | META-N | META-N-Studies | CHR | BP (GRCh37) | EA | NEA | EAF |
| --- | --- | --- | --- | --- | --- | --- | --- | --- | --- | --- | --- | --- |
| rs2814778 | 0.43 | 0.02 | 2.66E-87 | 0.28 | 0 | 5,793 | 6 | 1 | 159174683 | T | C | 0.036 |
| rs144109344 | -0.12 | 0.02 | 3.12E-10 | -0.06 | -0.06 | 5,976 | 7 | 2 | 136787730 | C | T | 0.964 |
| rs530475031 | 0.73 | 0.12 | 3.16E-09 | 0.47 | 0.46 | 4,952 | 5 | 12 | 48810860 | G | T | 0.998 |
| rs12747038 | -0.22 | 0.04 | 3.89E-09 | -0.13 | -0.08 | 5,976 | 7 | 1 | 146651428 | T | G | 0.990 |
| rs527921556 | 0.4 | 0.07 | 4.48E-09 | 0.33 | 0.31 | 5,793 | 6 | 6 | 160605701 | T | C | 0.996 |
| rs557482905 | 0.55 | 0.1 | 5.79E-09 | 0.42 | 0.38 | 3,778 | 4 | 5 | 80629499 | C | T | 0.998 |
| rs140048432 | -0.33 | 0.06 | 1.11E-08 | -0.25 | -0.25 | 5,976 | 7 | 9 | 17700893 | T | C | 0.996 |
| rs183362544 | 0.61 | 0.11 | 1.27E-08 | 0.28 | 0.28 | 2,717 | 4 | 2 | 97045902 | C | T | 0.998 |
| rs558204720 | 0.52 | 0.09 | 1.67E-08 | 0.37 | 0.33 | 1,486 | 2 | 16 | 59472815 | T | C | 0.998 |
| rs28734019 | -0.65 | 0.12 | 2.89E-08 | -0.53 | -0.5 | 4,124 | 4 | 1 | 90800573 | C | T | 0.998 |

The effect sizes of the primary GWAS index SNPs were compared with those from the SNPTEST/META GWAS. The direction was consistent and effect sizes were similar between the three GWAS, with those generated from the BOLT-LMM run (primary GWAS) being slightly larger, most likely due to the improved sample size (minor allele count) and power of the linear-mixed model (**Figure 4**). As expected, the META-WD effect size for the rs2814778 SNP was zero.

**Figure 4.**
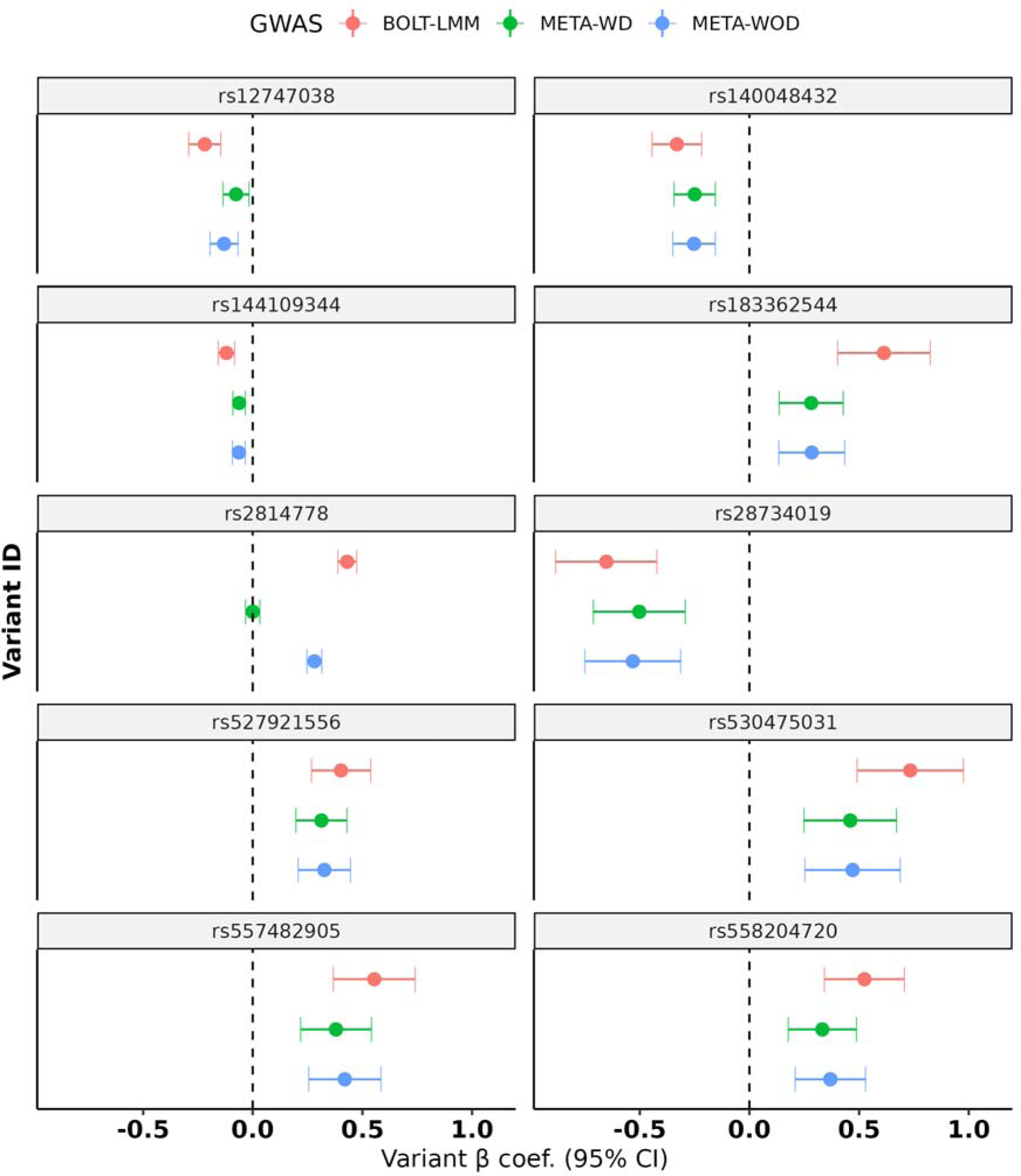
Effect estimates of the index SNPs. The beta coefficient for each index SNP is displayed along with 95% CIs. These are displayed for the BOLT-LMM, META-WOD and META-WD GWAS.

We next investigated the association statistics of the index SNPs in each Kpop. This was done to detect discrepancies in directionality and effect sizes, which could indicate residual population structure or a SNP association with a specific Kpop. Overall, there was agreement in direction, and some variation in effect sizes was detected across Kpops (**Supplementary Figure 5).**

The GCTA-COJO analysis was also run on the two SNPTEST/META GWASs. The META- WOD analysis identified rs2814778, rs138163369 and rs570518709 as index SNPs. Similarly, the META-WD analysis identified rs138163369 and rs570518709. These two latter SNPs were not identified as index SNPs in the BOLT-LMM analysis, but their P-values were similar (rs138163369 – 4.90E-08, 2.28E-08, 1.22E-08; rs570518709 – 8.10E-08, 1.07E-09, 3.03E-09) (**Supplementary Table 2**). As another sensitivity analysis to test the reliability of the BOLT-LMM results, the effect sizes of all GCTA-COJO SNPs were compared in a pair-wise manner across the three GWAS. A regression line was fit through the scatter plots, showing a large degree of correlation between the BOLT-LMM effect sizes and the SNPTEST/META runs (META-WOD R^2^ = 0.91, META-WD R^2^ = 0.93) (**Supplementary Figure 6).**

Two PLINK clumping analyses were performed on the filtered AFR_CAG summary statistics using the same clumping parameters on the well-known FUMA platform [43]. Here, 193 SNPs were identified as loci at the relaxed threshold of r^2^=0.6 and 73 independent loci at the stringent threshold of r^2^=0.1. Finally, 12 top loci were identified at r^2^=0.001 and a 10Mb window, which are the very conservative MR clumping parameters [44,45]. Furthermore, a FUMA analysis was run on the filtered AFR_CAG dataset for the top loci (r^2^=0.1). This was done to visualise which genomic locations are affecting neutrophil count and if they are more likely to have a particular genetic function compared to the whole genome i.e. functional variants [70]. Seventeen genomic risk loci were identified (**Supplementary Figure 7A**). The ANNOVAR analysis [71] showed evidence for changes in genetic function enrichment relative to all SNPs in the reference panel. In brief, seven genomic regions were enriched, all indicating an enrichment in genic rather than intergenic spaces (**Supplementary Figure 7B**).

Next, we investigated the independent SNPs in the GWAS Catalog [72], as we aimed to see if they have been previously associated with WBC count or immunity. Here, SNPs predominantly showed associations with white blood cell count variation, further improving the reliability of the GWAS (**Supplementary Table 3**). We compared the AFR_CAG GWAS with a neutrophil count GWAS meta-analysis of Africans from UKBB and additional studies from Chen et al. [33], and found that 81.71% of the GWAS significant SNPs from Chen et al. were replicated (using the same covariates) in the AFR_CAG dataset (P<0.05) (**Supplementary Table 4**). The Manhattan plots also visually showed a good degree of overlap (**Supplementary Figure 8**), in contrast with a GWAS of neutrophil count in Europeans **Supplementary Figure 9**) [38]. Finally, SNPs that were top loci at r^2^=0.1 were investigated in the Astle et al. [38] and Chen et al. [33] summary statistics, as well as in the GWAS Catalog [72]. Nineteen genetic variants were not present in these three datasets, 7 of which were index SNPs (**Table 3**).

**Table 3.** Top loci not found in other studies. Only independent SNPs clumped at r = 0.1 are shown.

| SNP | CHR | BP (GRCh37) | r0.001 lead? | cojo_index | Novel? | Nearest gene | Type |
| --- | --- | --- | --- | --- | --- | --- | --- |
| rs28734019 | 1 | 90800573 | Yes | Yes | Yes | RNU6-695P | Intergenic |
| rs61823703 | 1 | 159542164 | No | No | Yes | OR10AE1P | Intergenic |
| rs539456851 | 1 | 158731459 | No | No | Yes | OR6N1 | Intergenic |
| rs371178711 | 1 | 158186653 | No | No | Yes | RP11-404O13.5 | Intergenic |
| rs146677619 | 1 | 158995984 | No | No | Yes | IFI16 | Intronic |
| rs11576058 | 1 | 161111446 | No | No | Yes | UFC1 | Intergenic |
| 1:158777618_CT_C | 1 | 158777618 | No | No | Yes | OR10AA1P | Downstream |
| rs183362544 | 2 | 97045902 | Yes | Yes | Yes | NCAPH | Intergenic |
| rs11422063 | 1 | 159799599 | No | No | Yes | SLAMF8 | Intronic |
| rs112483667 | 1 | 151651180 | No | No | Yes | SNX27 | Intronic |
| rs12406899 | 1 | 157540651 | No | No | Yes | FCRL4 | Intergenic |
| rs1103805 | 1 | 158924741 | No | No | Yes | PYHIN1 | Intronic |
| rs557482905 | 5 | 80629499 | Yes | Yes | Yes | ACOT12 | Intronic |
| rs527921556 | 6 | 160605701 | Yes | Yes | Yes | SLC22A2 | Intronic |
| rs10096834 | 8 | 116281087 | Yes | No | Yes | TRPS1 | Intergenic |
| rs140048432 | 9 | 17700893 | Yes | Yes | Yes | SH3GL2 | Intronic |
| rs530475031 | 12 | 48810860 | Yes | Yes | Yes | C12orf54 | Intronic |
| rs558204720 | 16 | 59472815 | Yes | Yes | Yes | LOC105371298 | Intronic |
| rs138163369 | 18 | 6492075 | Yes | No | Yes | CTD-2124B20.2 | Intergenic |

### Heritability analysis

Without adjusting for rs2814778, the genetic variance was estimated at 0.101 (10.1%) (SE = 0.018), and the phenotypic variance at 0.133 (13.3%) (SE = 0.003) with an analysis P-value of 2.29e-09. When adjusting for the ACKR1/Duffy SNP, the genetic variance was estimated at 0.050 (5%) (SE = 0.017), twice as low as in the previous analysis, and the phenotypic variance was estimated at 0.123 (12.3%) (SE = 0.002), with the analysis P-value of 1.36E-03 (Supplementary Table 5).

### Descriptive analyses of neutrophil count

Next, we aimed to assess if the index SNPs remained associated with neutrophil count when adjusting for variables such as BMI and smoking status. This was done to investigate the reliability of the index SNPs in the context of their relationship with neutrophil count. First, the descriptive statistics of the AFR_CAG dataset were studied with these additional variables (**Supplementary Table 6**). Several variables had missing data or had values assigned as “prefer not to answer” / “not sure” in the case of self-reported traits. There was no evidence of a difference in neutrophil count between these data types and those that were kept in the dataset (**Supplementary Table 7**). 5,310 individuals remained in the dataset after filtering out these data types. The proportion of variance explained for the additional covariates was explored with an ANOVA analysis (**Supplementary Figure 10**).

### BOLT-LMM with additional covariates

A sensitivity BOLT-LMM GWAS was conducted with six additional covariates on 5,310 individuals: UN region of birth, K-means cluster, smoking status, alcohol drinker status, menstrual status and BMI. The association statistics of this sensitivity run and the main BOLT-LMM GWAS run were compared, showing very similar results (**Supplementary Table 8**). This provides evidence that the effect of these additional variables on the main GWAS were modest, and that the PCs and kinship matrix derived by BOLT-LMM appears to have accounted for any population stratification.

### Mendelian randomization

Finally, a bi-directional MR was performed between neutrophil count and severe malaria. For the latter, we used summary statistics from the MalariaGEN study [37]. Only 3 SNPs were available to proxy for neutrophil count after data harmonization with the malaria dataset. For severe malaria as an exposure, 7 SNPs were available for overall severe malaria, 2 for CM and 3 for OTHER.

The MR analysis did not suggest an effect of increasing neutrophil count on CM risk (IVW OR: 1.00, 95% CI: 0.94 to 1.06; P = 0.98. There was limited evidence of an effect of neutrophil count on overall severe malaria (IVW OR: 1.03, 95% CI: 0.98 to 1.07; P = 0.24), OTHER (IVW OR: 1.03, 95% CI: 0.98 to 1.09; P = 0.26) and SMA (IVW OR: 1.08, 95% CI: 0.99 to 1.18; P = 0.08), although the effect estimates were trending towards an increased risk of severity, particularly for SMA (**Figure 5A, Supplementary Table 9**). When running the MR analysis in the other direction, there was little evidence of an effect of overall severe malaria (IVW OR: 2.03, 95% CI: 0.70 to 5.84; P = 0.19), CM (IVW OR: 2.14, 95% CI: 0.70 to 6.57; P = 0.18) and OTHER (IVW OR: 2.08, 95% CI: 0.59 to 7.34; P = 0.25) on neutrophil count. However, there was a directional agreement in effect estimates towards an increase in neutrophil count (**Figure 5B, Supplementary Table 9**). No SNPs passed the GWAS significance threshold for SMA, meaning this analysis could not be conducted.

**Figure 5.**
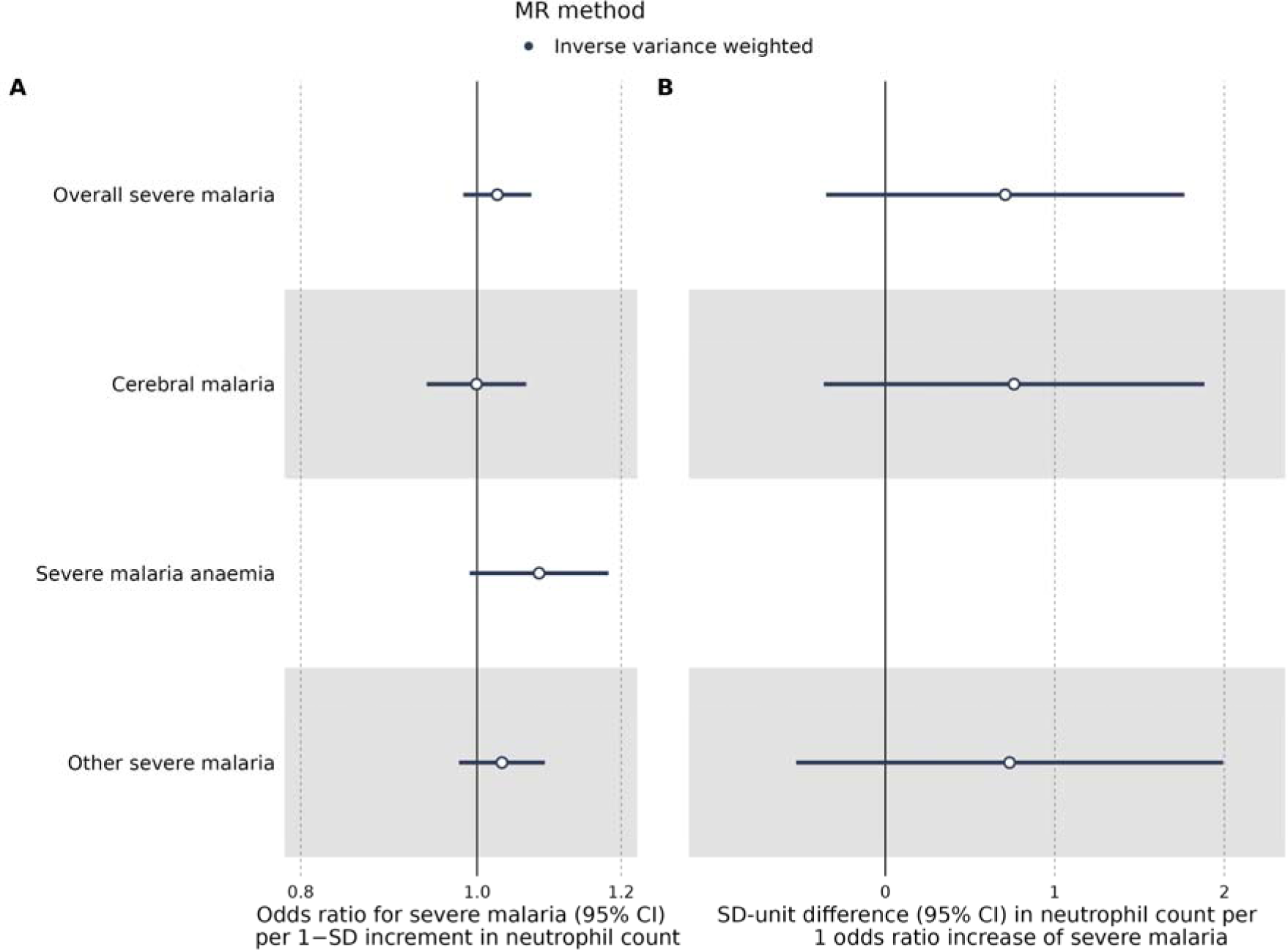
Bi-directional Mendelian randomization. Forest plot of the IVW MR analysis with neutrophil count as an exposure (**A**) and severe malaria as an exposure (**B**). Overall severe malaria and its sub-phenotypes are listed on the y-axis, with the effect estimates on the x-axis. In the first instance, the MR results are interpreted as an OR increase severe malaria per 1-SD increase in neutrophil count, while in the latter as a 1-SD unit difference in neutrophil count per 1-OR

A single-SNP MR analysis was performed to study the effect of each genetic variant on the outcome. For neutrophil count as the exposure, SNPs rs2325919 (proxy for rs2814778), rs7460611 (proxy for rs10096834), and rs144109344 were used. There was little evidence of an effect by any single SNP, although the general direction was towards an increased risk of severe malaria (**Supplementary Table 10**). The estimated conditional F-statistic for SNPs rs2325919, rs7460611 and rs144109344 were 182, 16 and 36 respectively. For severe malaria as an exposure, SNPs rs113892119, rs116423146, rs1419114, rs553707144, rs557568961, rs57032711, rs8176751 were used to proxy for overall severe malaria, rs113892119 and rs543034558 for CM, and rs113892119, rs116423146, rs557568961 for OTHER (**Supplementary Table 11**). The estimated conditional F-statistic for SNPs rs113892119, rs116423146, rs1419114, rs553707144, rs557568961, rs57032711 and rs8176751 were 96, 32, 30, 38, 119, 32 and 44 respectively.

## Discussion

Here, we conducted a GWAS of neutrophil count in individuals from the AFR CAG in UKBB. Seventy-three independent loci were identified, of which nineteen were novel and rare. Ten index SNPs were found using the conservative GCTA-COJO approach, and another two through MR clumping. Moreover, BOLT-LMM was found to be reliable in conducting GWAS on UKBB participants of African ancestry. Ultimately, this allowed us to run an MR analysis between neutrophil count and *P. falciparum* severe malaria.

An aim of our study was to assess whether BOLT-LMM could provide reliable results when performing a GWAS in people of non-European ancestry, such as those in the UKBB AFR CAG. In their meta-analysis of BCT in non-European datasets, Chen at al. used a linear model in PLINK to run their GWAS, restricting BOLT-LMM only to the European dataset [33]. Compared to our META-WD and META-WOD GWAS, the BOLT-LMM approach was more similar to that of Chen et al conducted with a larger sample-size (N=15,171). These findings indicate that a linear mixed model framework using a kinship matrix might reliably account for extensive population structure in a complex data set such as that seen in the African CAG used here. If this observation holds true this would be advantageous in identifying more causal ancestry-specific SNPs in future studies, as the power of BOLT-LMM scales with increasing GWAS sample-size [67].

Next, we found a marked difference between the genetic architecture of neutrophil count in people of African vs. European ancestry [38]. Interestingly, tissue expression for BCTs has been found to vary between ancestries as well [73], further showing the importance of conducting GWAS in diverse populations to improve the understanding of BCT biology. We investigated some of the GCTA-COJO index SNPs in relation to a biological mechanism that could explain how allele variation might affect neutrophil count levels in people of African ancestry.

One such SNP is rs12747038, an index SNP located on chromosome 1 (1q21.1), was also identified by Chen et al. and Hu et al. to be associated with neutrophil count and they found a similar effect size to us (AFR_CAG: BETA = -0.22, P-value = 3.90e-09; Chen et al: BETA = -0.31, P-value = 3e-20; Hu et al: BETA = -0.21, P-value = 8e-36) [33,74]. Interestingly, rs12747038 has a role as a splicing QTL (sQTL) i.e. affecting alternative splicing to make different protein isoforms [75], which can be more relevant mechanistically to a phenotype compared to expression data [76]. The strongest association as an sQTL was with *NBPF12* gene (NES = 0.49, P-value = 2.9e-9) in the thyroid. McCartney et al. had found that rs11239931, an sQTL for *NBPF12*, was also associated with a decrease in granulocyte count (BETA = -0.23, P-value = 4e-12) in people of African ancestry (N=6,152) [77]. *NBPF12* is part of the neuroblastoma breakpoint family, which has been associated with an array of traits, such as autism, psoriasis and various cancers [78].

The rs2814778 (chromosome 1q23.2) index SNP has been the most replicated genetic variant in people of African ancestry known to affect neutrophil count [33,79–84], with the CC genotype (most common in Africans) associated with decreased neutrophil count [20]. The exact location of rs2814778 is inside a promoter upstream of the *ACKR1*/*DARC* (Atypical Chemokine Receptor 1/Duffy Antigen Receptor for Chemokines) gene [13]. The CC genotype inhibits the binding of the GATA transcription factor and therefore *ACKR1* expression in erythrocytes, preventing the production of a glycosylated transmembrane receptor [20]. This receptor is heavily involved in chemokine signalling, such as CXCL8 and CCL5 [13].

rs144109344 is an index variant on chromosome 2 (2q21.3), and its association was similar to that in the studies of Chen et al. and Soremekun et al. (N=17,802 Africans): AFR_CAG BETA = -0.12, P-value = 3.10e-10; Chen BETA = -0.27, P-value = 3.39e-14; Soremekun BETA = -0.21, P-value = 2e-13) [33,82]. Similarly, other SNPs mapping to the *DARS*/*CXCR4* (Aspartyl-TRNA Synthetase 1/C-X-C Motif Chemokine Receptor 4) genes have been associated with neutrophil and monocyte count [33,38,85–88]. CXCR4 is a chemokine receptor which binds to CXCL12 [89], and is known to regulate the release of neutrophils from the bone marrow during both homeostasis and infections [90]. Interestingly, CXCR4 has been implicated in *P. falciparum* pathogenesis. Macrophage migration inhibitory factor (MIF) can interact with CXCR4 to recruit neutrophils [91], and *P. falciparum* is known to also produce MIF (PfMIF) [92]. A previous laboratory study using both murine (*P berghei*) and human (*P falciparum*) models found impairment of the parasite liver-cycle in both genetically deficient and drug-targeted CXCR4 [93].

We note that the process of mapping SNPs to a biological function is a difficult process, and the brief discussion above only serves as an inquiry into a possible explanation for the primary GWAS results.

Finally, in the MR analysis, there was limited evidence for an effect of increased circulating neutrophil on the risk of SM. The strongest effect was observed for the SMA sub-phenotype, however, this did not reach statistical significance. Interestingly, a recent report demonstrated an association between circulating neutrophil transcriptional activity and levels of anaemia in children with malaria [24], highlighting the need for further pathophysiological studies. We also observed little evidence for an effect of SM on neutrophil count. Previously, Band et al. performed a MR analysis between neutrophil count and *P. falciparum* SM [37], however, they used SNPs for neutrophil count generated from a GWAS in Europeans from UKBB [38], where they found no evidence of an effect on SM (AFR_CAG BETA = 0.03, P-value = 0.24; Band BETA = 0.00, P-value = 0.87) [37].

Our study has certain limitations. Firstly, the novel genetic variants identified here may be a result of Winner’s curse [94] - SNPs can pass the “significance” threshold (commonly set at 5e-8 [95,96]) in GWAS by chance in the first discovery study, which is then not replicated in subsequent studies [97,98].

Secondly, only a limited number of instruments were available to proxy for neutrophil count in the MR analysis. Seven index SNPs had a very high effect allele count, which might have been fixed in the MalariaGEN study population and so could not be used in the MR analysis. The rs2814778 SNP (associated with the *ACKR1* gene) most likely had a very small allele frequency and might have been eliminated, although we were able to use another SNP in LD with it as a proxy. While LD proxies are useful, they can also come with the caveat of not precisely instrumenting the trait [36].

Finally, the most impactful limitation in this study is the small sample-size and hence statistical power. As mentioned previously, we have chosen to use BOLT-LMM here to best address the issues of a small sample-size and the presence of population structure. Current studies performed on people living in sub-Saharan Africa have been small [33,80–82] compared to those currently being carried out in Europe, East Asia and the US [31,88,99]. Having a large-scale study akin to UKBB in sub-Saharan African would allow for finding common SNPs with smaller effect sizes that could be used reliably for polygenic risk score generation or MR analyses for complex traits such as neutrophil count.

In conclusion, our GWAS of neutrophil count in people from the UKBB African CAG identified several SNPs associated with neutrophil count. Additionally, our analyses would support a conclusion that linear mixed model frameworks can properly account for possible confounding due to population stratification in complex highly stratified sample populations. Finally, while the MR results were largely inconclusive, this only demonstrates the importance of conducting large-scale biobank studies in Africa.

## Availability of data and materials

Genetic data from UK Biobank were made available as part of project code 15825. Analytical code is available on GitHub at https://github.com/andrewcon/AFR-GWAS-neutrophil.

## Contributions

AC, BA, DA, EEV, CJB and REM conceived the study. AC conducted the analysis. All authors contributed to the interpretation of the findings. AC, EEV, CJB, DH and BA wrote the manuscript. All authors critically revised the paper for intellectual content and approved the final version of the manuscript.

## Supporting information

Suppementary Methods

STROBE-MR

## Acknowledgements

We are grateful to the UK Biobank study and its participants. This research has been conducted using the UK Biobank resource under Application 15825. We thank the Malaria GEN Network for their study and their participants.

## Ethics declarations

### Ethics approval and consent to participate

UK Biobank received ethical approval from the NHS National Research Ethics Service North West (11/NW/0382; 16/NW/0274) and was conducted in accordance with the Declaration of Helsinki. All participants provided written informed consent before enrolment in the study.

### Consent for publication

All authors consented to the publication of this work.

## Supplementary information

Additional file 1 Supplementary Figures

Additional file 2 Supplementary Tables

Additional file 3 Supplementary Methods

Additional file 4 STROBE-MR checklist

## SUPPLEMENTARY FIGURES

**Figure S1.**
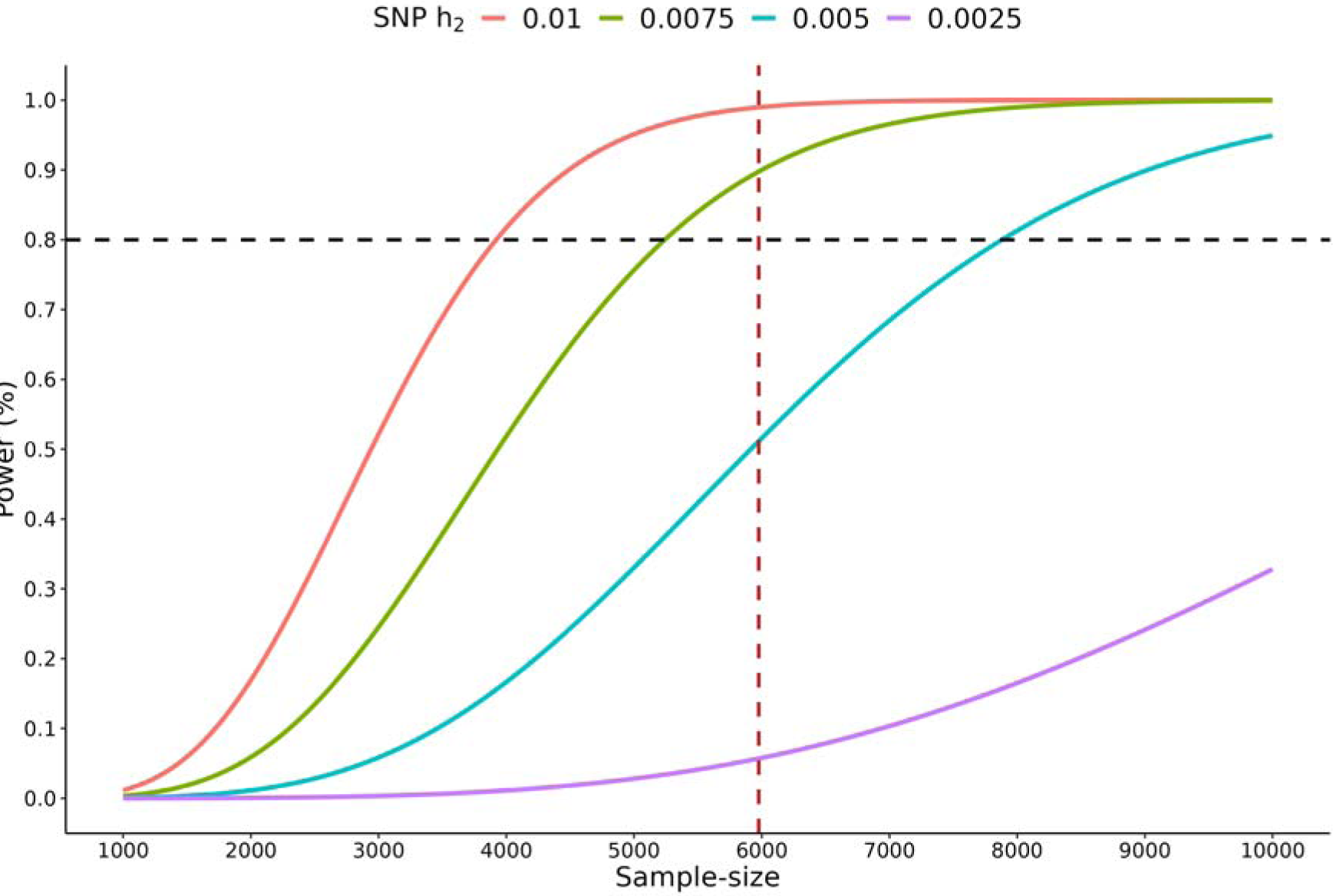
Power calculation of a GWAS AFR_CAG sample. The x-axis indicates the sample-size, while the y-axis is the statistical power of an association test. Each curved line shows how power varies by sample-size at different degrees of the variance explained by all the SNPs on neutrophil count (1%, 0.75%, 0.5%, 0.25%). A black horizontal line is fixed at Power=80%, and a red vertical line is drawn at the GWAS sample size of 5,976.

**Figure S2.**
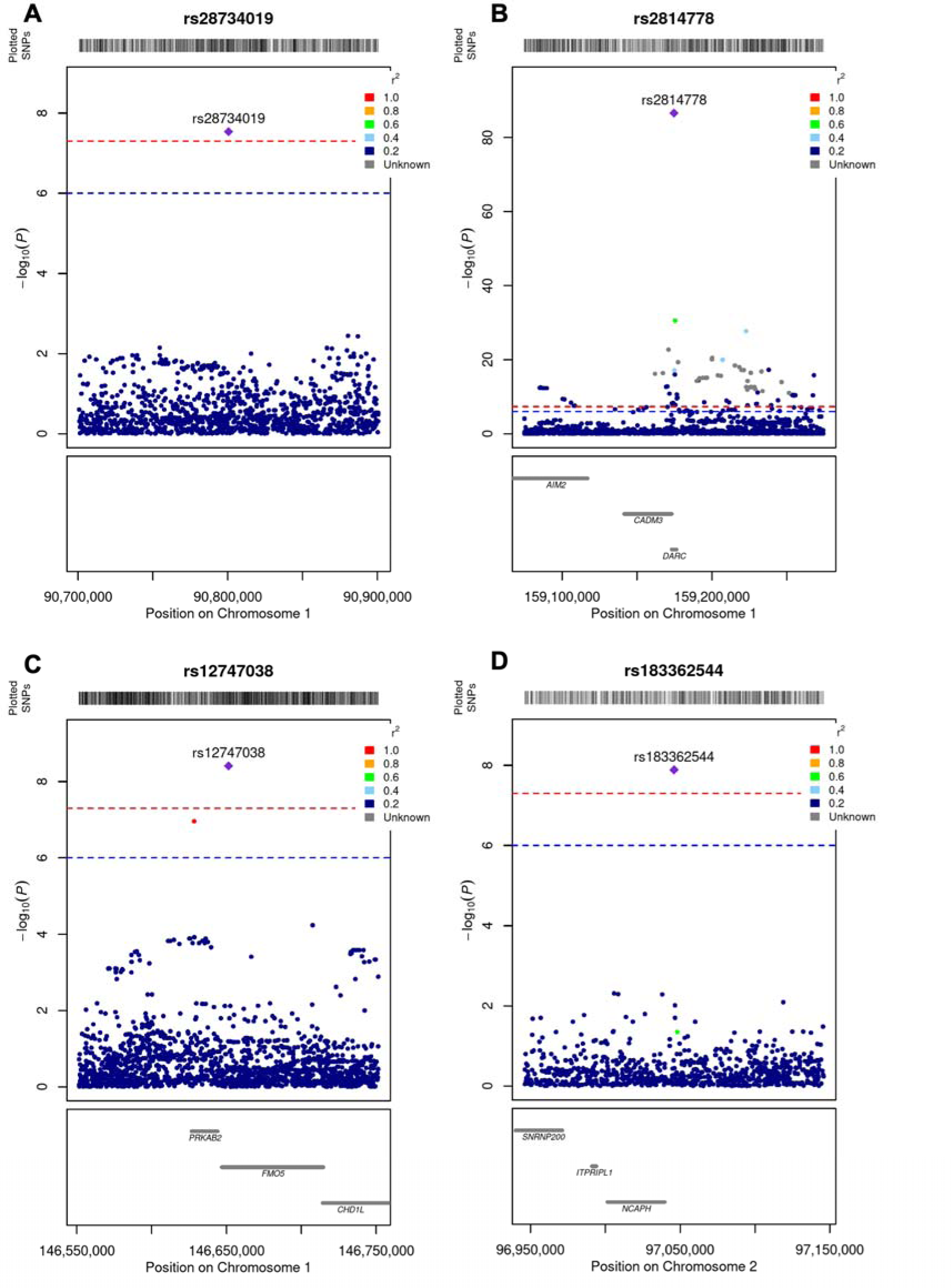
Regional plots of index SNPs (1).

**Figure S3.**
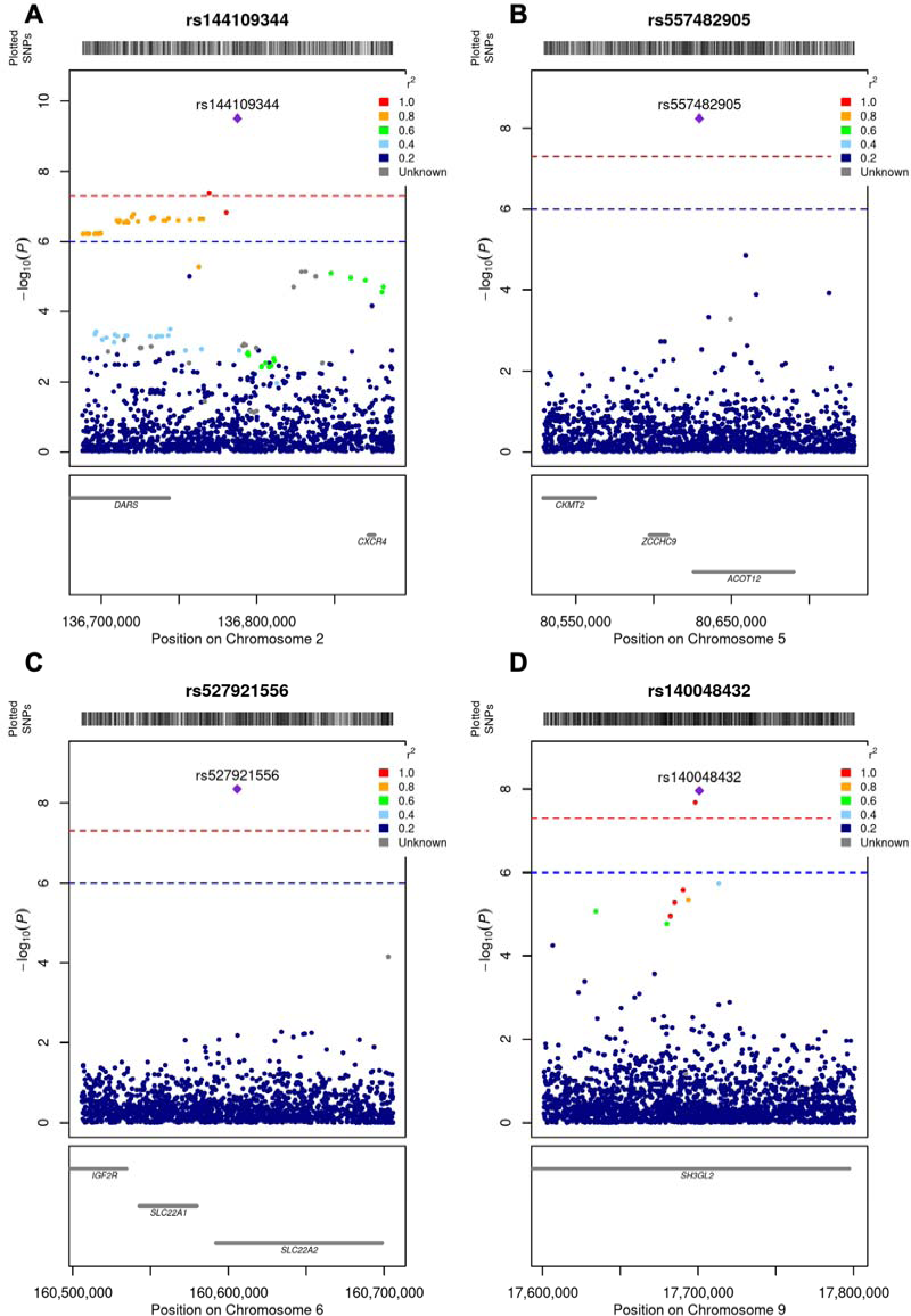
Regional plots of index SNPs (2).

**Figure S4.**
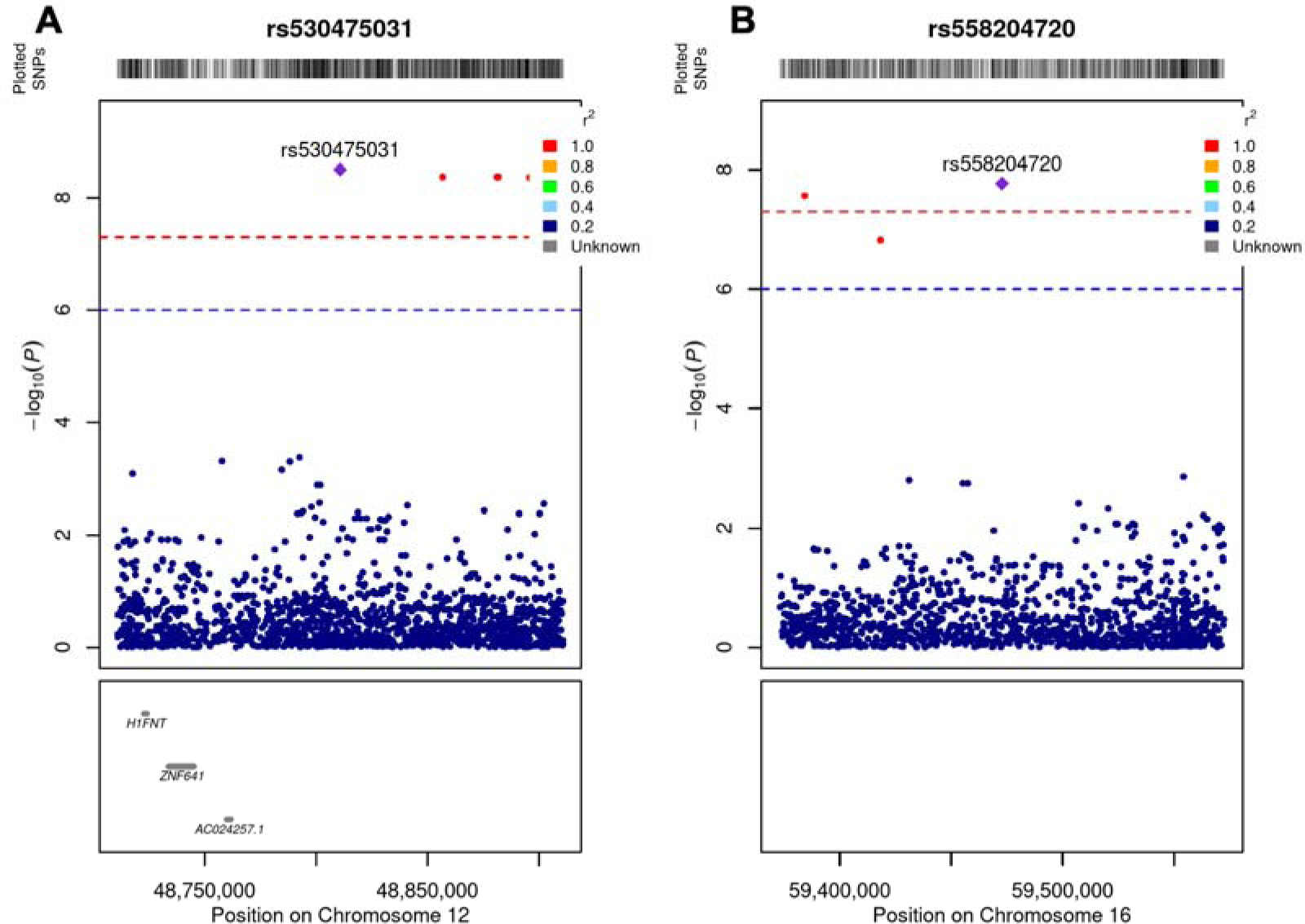
Regional plots of index SNPs (3).

**Figure S5.**
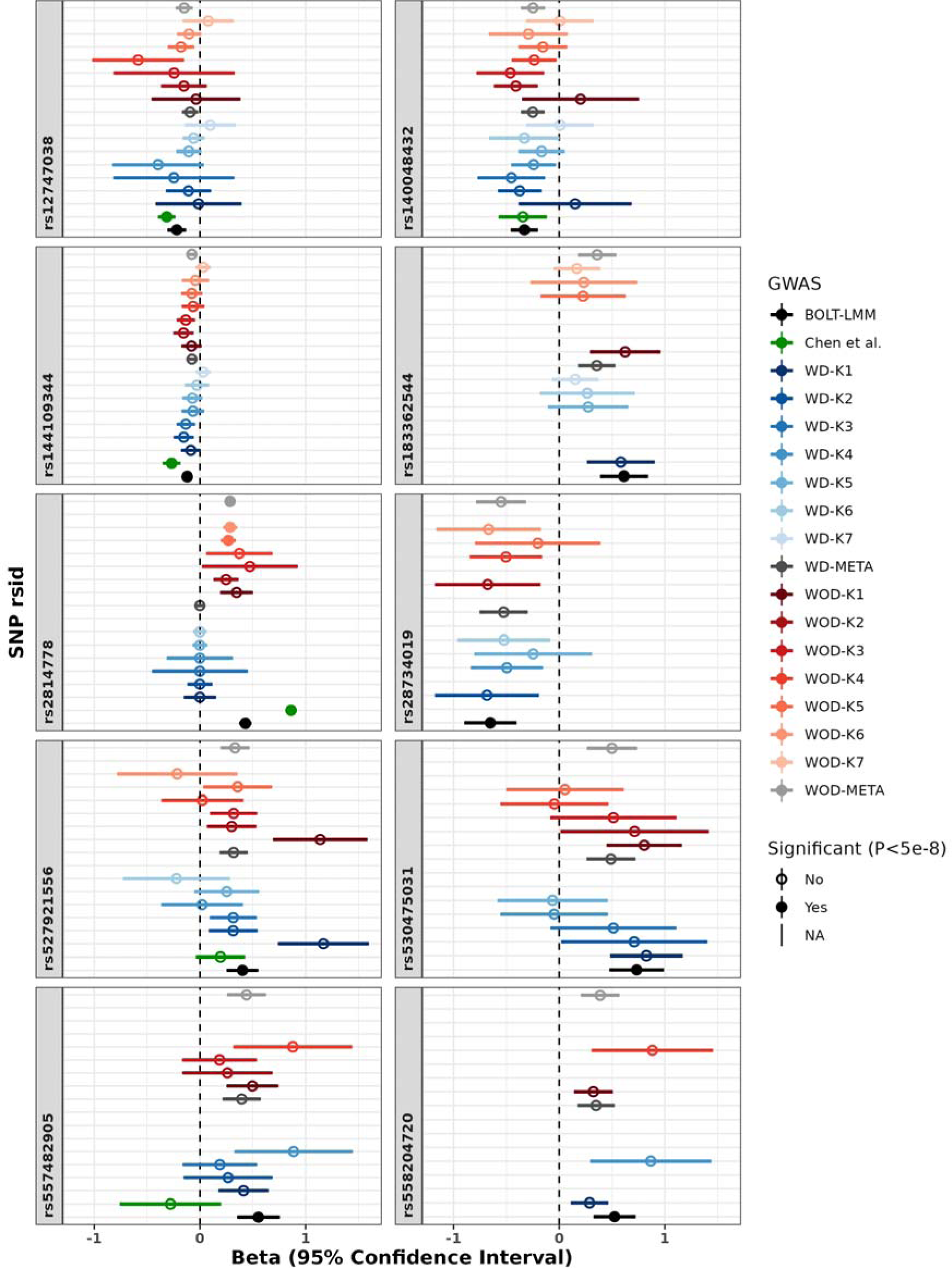
Forest plot of index SNPs by K-means cluster. The effect-size of each BOLT index SNP was compared to that from SNPTEST/META, by-Kpop runs and Chen et al GWAS. Effect-sizes for each SNP across GWAS are present in the respective boxes of the figure. The x-axis indicates the effect-size (beta coefficient) of each SNP with 95% CIs, while the y-axis is the type of GWAS (indicated by the figure legend colouring). Some effect sizes were not displayed, either due to a low minor allele count in the case of the Kpop GWAS, or due to not being present in the summary statistics, in the case of the Chen GWAS. Non-signif. WD = adjusting for rs2814778; WOD = without adjusting for rs2814778.

**Figure S6.**
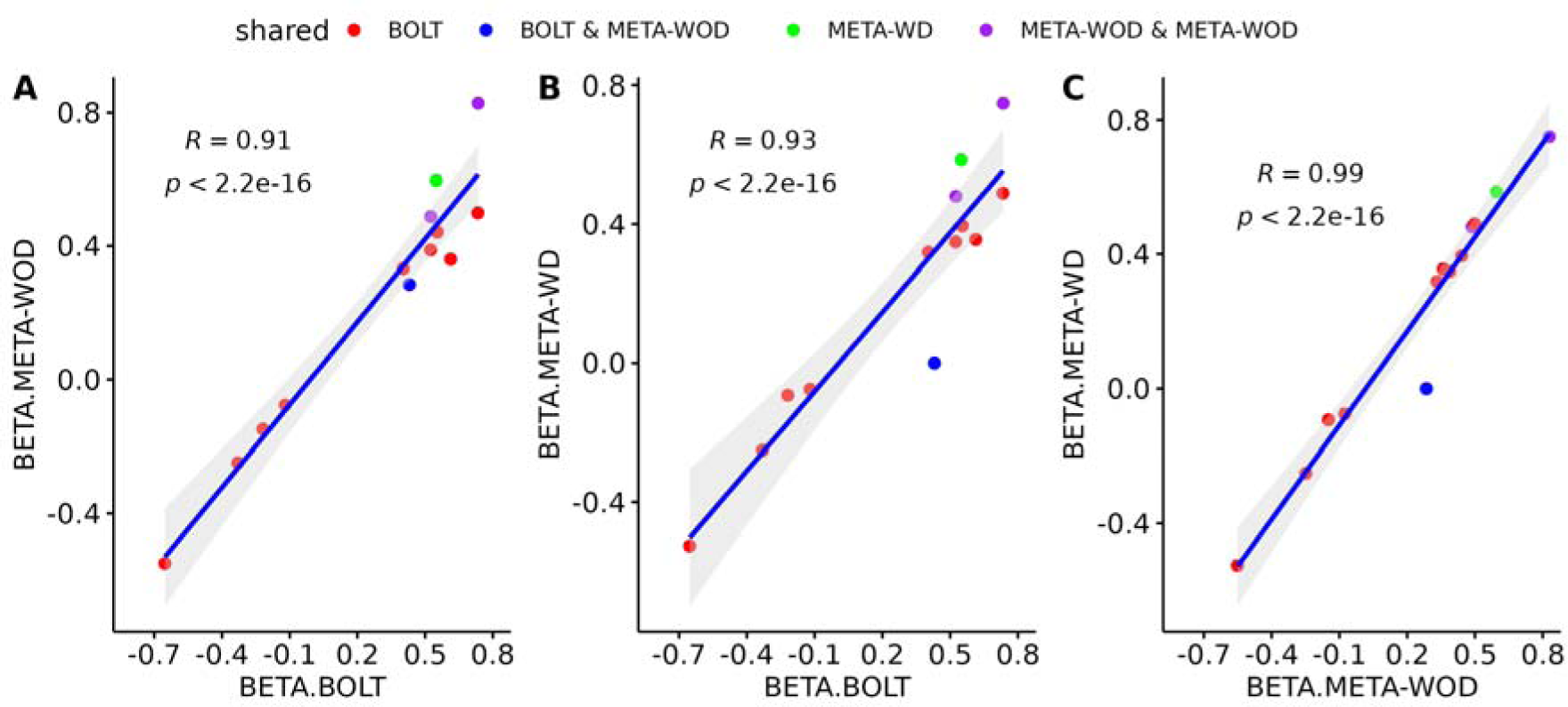
Scatter plot of GCTA-COJO effect sizes. Comparison of effect sizes of all GCTA-COJO independent signals of BOLT-LMM with META-WOD (A), BOLT-LMM with META-WD (B) and META-WOD with META-WD (C).

**Figure S7.**
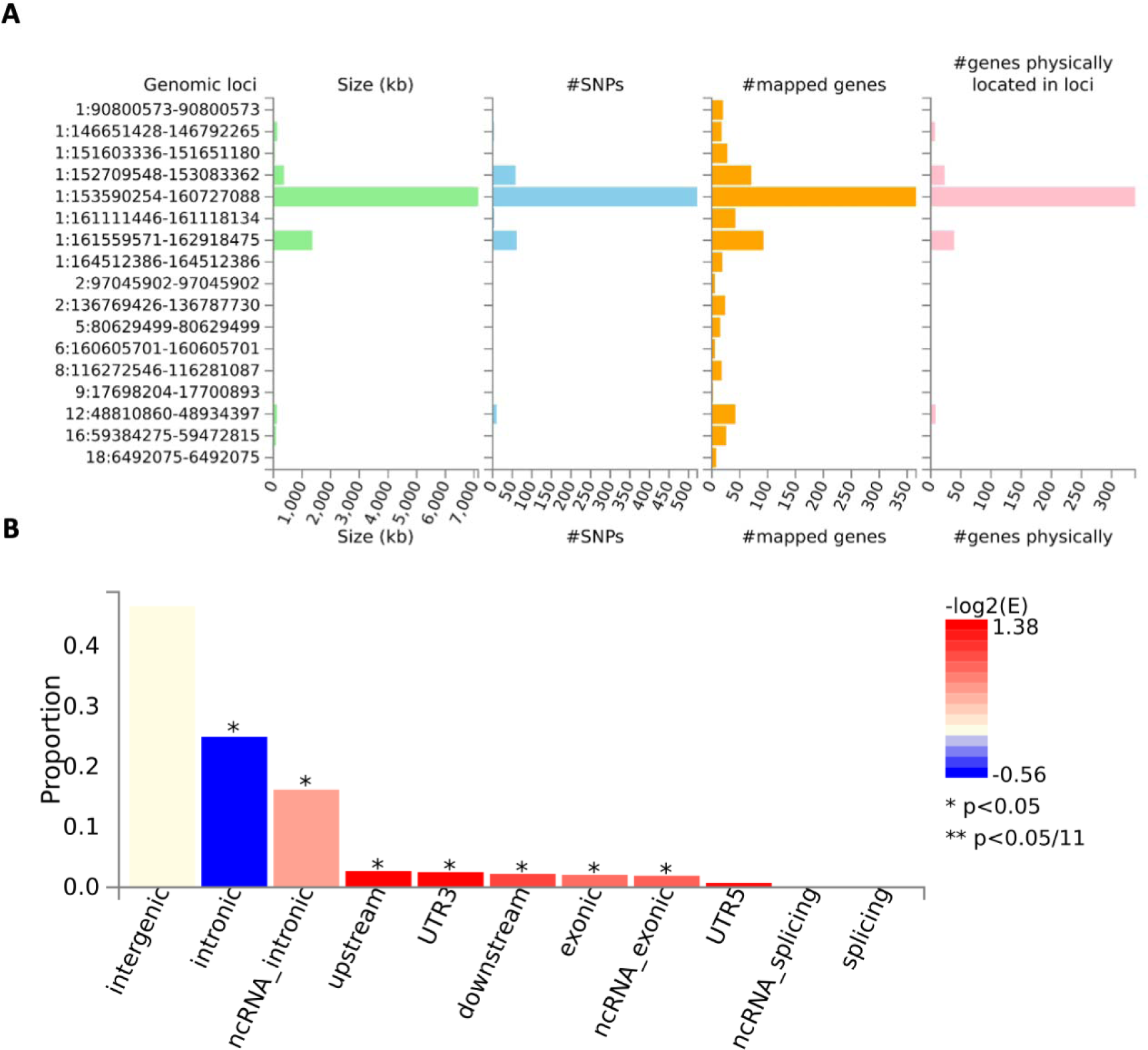
Description of genomic risk loci. FUMA analysis results for SNPs passing the GWAS significance threshold in the BOLT-LMM filtered GWAS.

**Figure S8.**
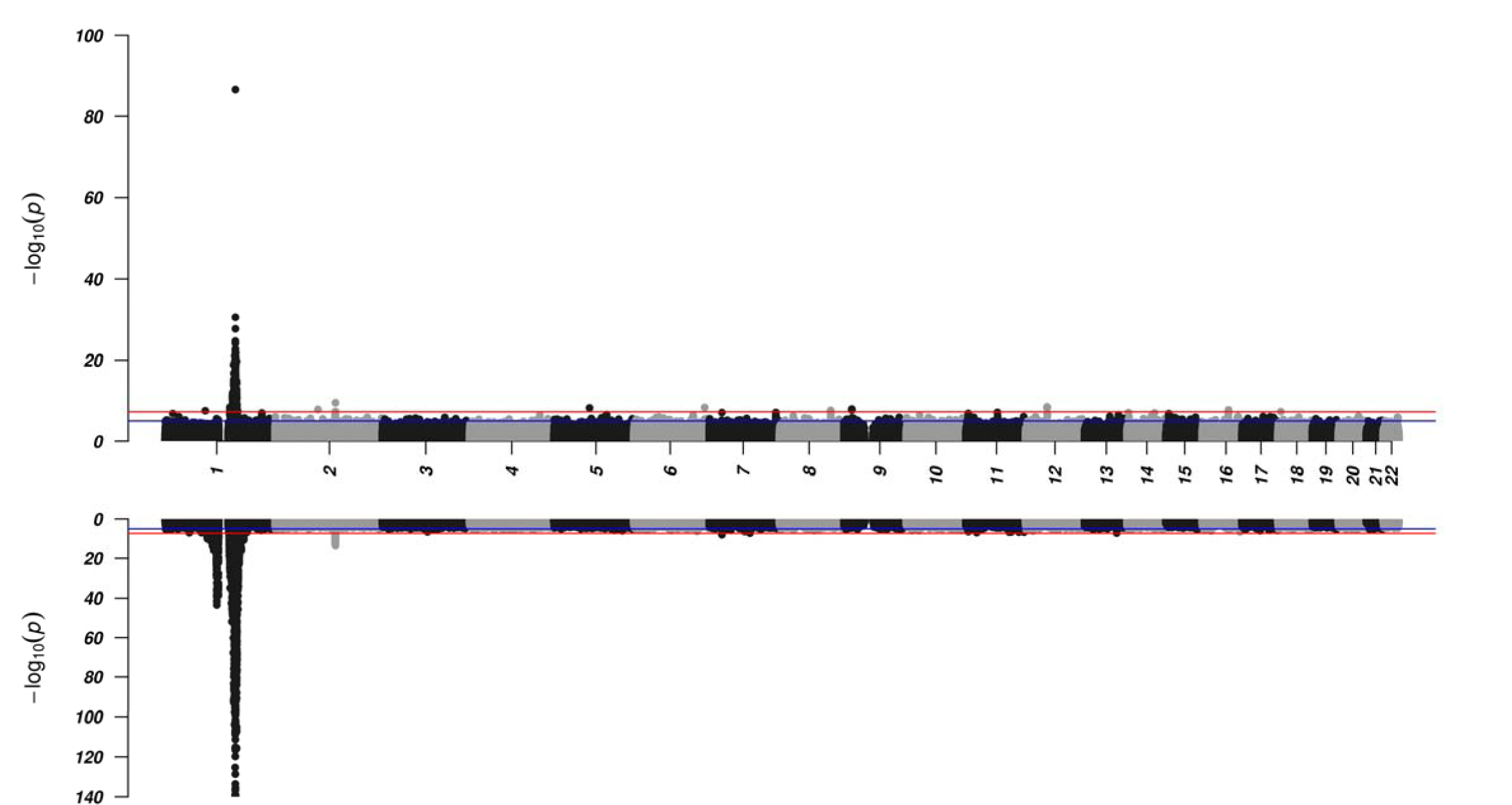
Comparison of GWAS results for neutrophil count in Africans. Manhattan plot of BOLT-LMM neutrophil count WAS from our study (top) mirrored with another Manhattan plot generated using summary statistics from a GWAS of utrophil count done in people of African ancestry (Chen et al, reference).

**Figure S9.**
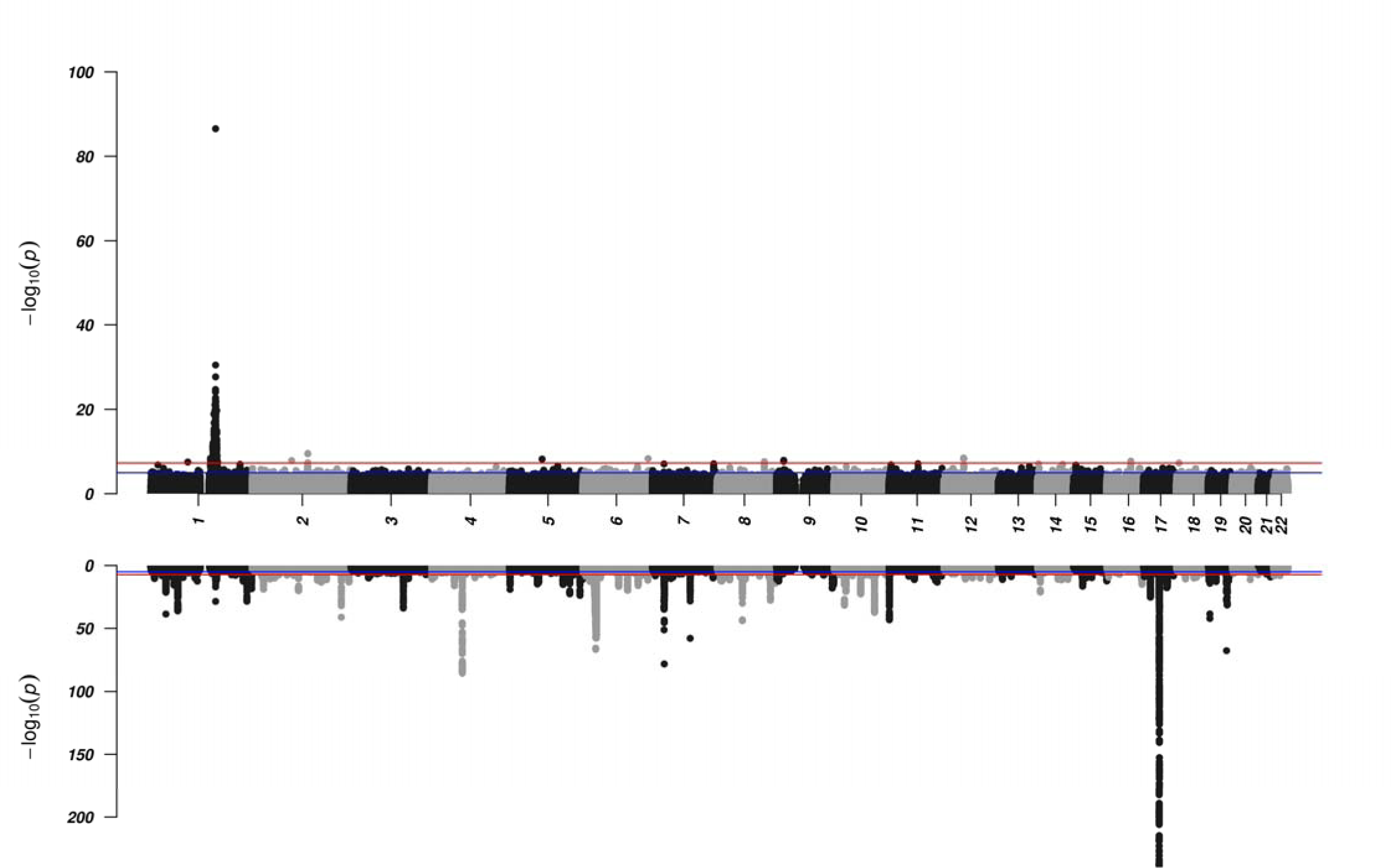
Comparison of GWAS results for neutrophil count in Europeans. Manhattan plot of BOLT-LMM neutrophil count GWAS from r study (top) mirrored with another Manhattan plot generated a GWAS of neutrophil count done in people of European ancestry in UK obank by Astle et al 10.1016/j.cell.2016.10.042.

**Figure S10:**
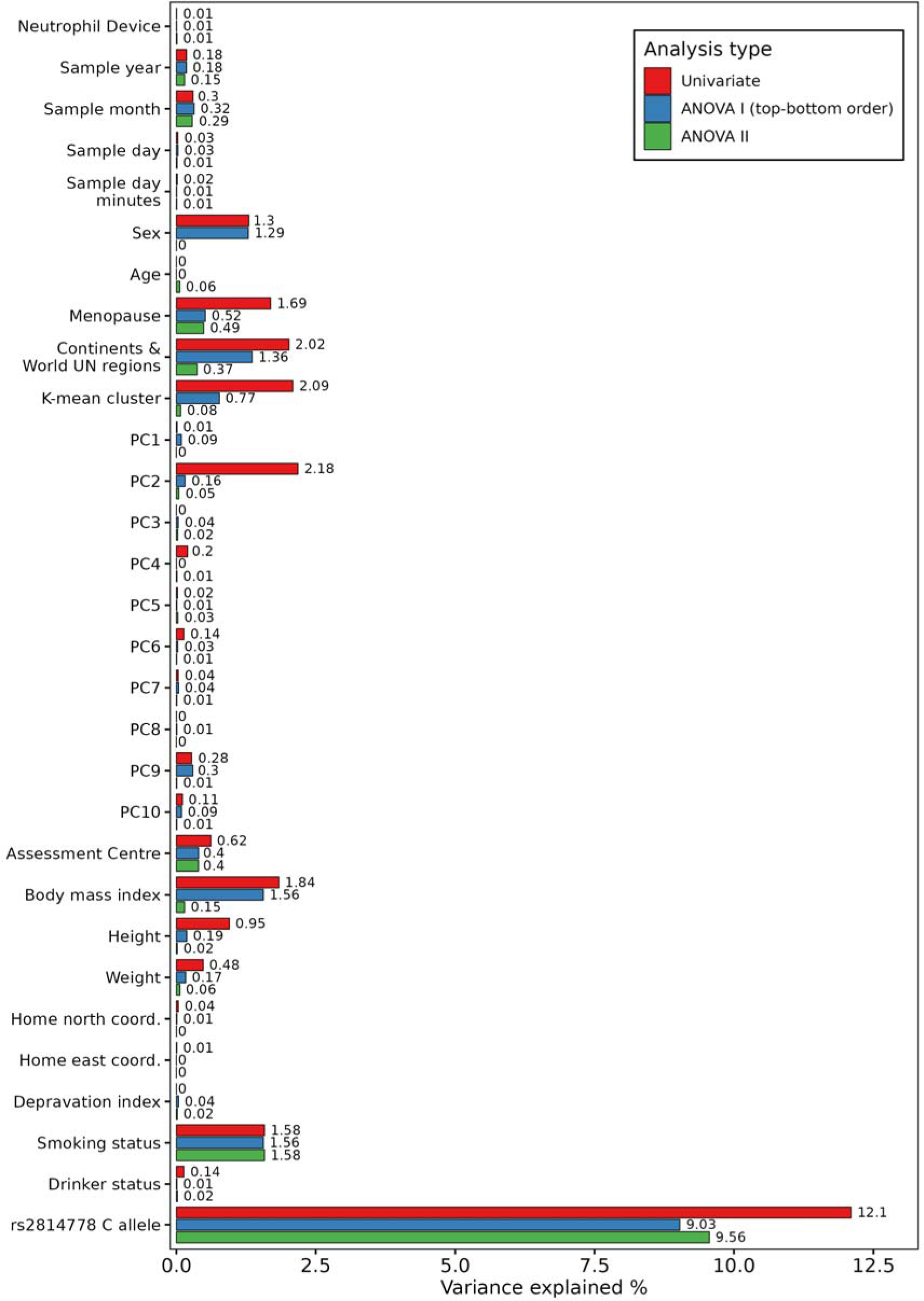
Proportion of variance explained by traits on neutrophil count. The x-axis indicates the trait studied, while the y-axis indicates the proportion of variance explained (PVE) on neutrophil count by that trait. The PVE of each trait was studied in a univariable manner (red), adjusted for only the other traits above in an ANOVA I hierarchical manner (blue), and adjusted between all other traits in a ANOVA II manner.

**Figure S11:**
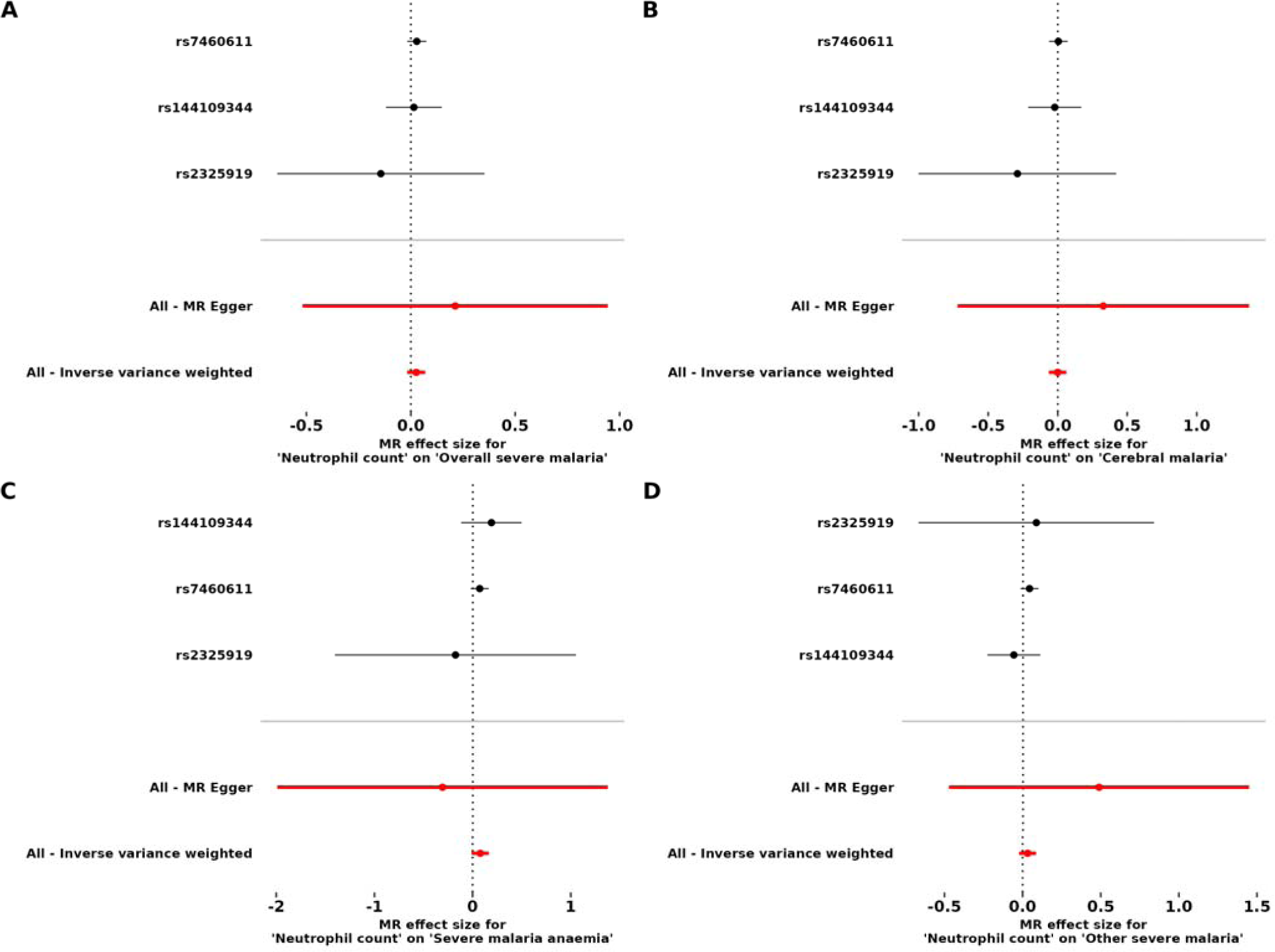
Single-SNP MR analysis of neutrophil count on severe malaria and its subtypes. SNPs proxying for neutrophil count are shown on the x-axis. The effect of each SNP proxying for neutrophil count is displayed on the y-axis, along with 95% CIs. The MR-Egger and IVW MR methods are shown below the single-SNP analysis.

**Figure S12:**
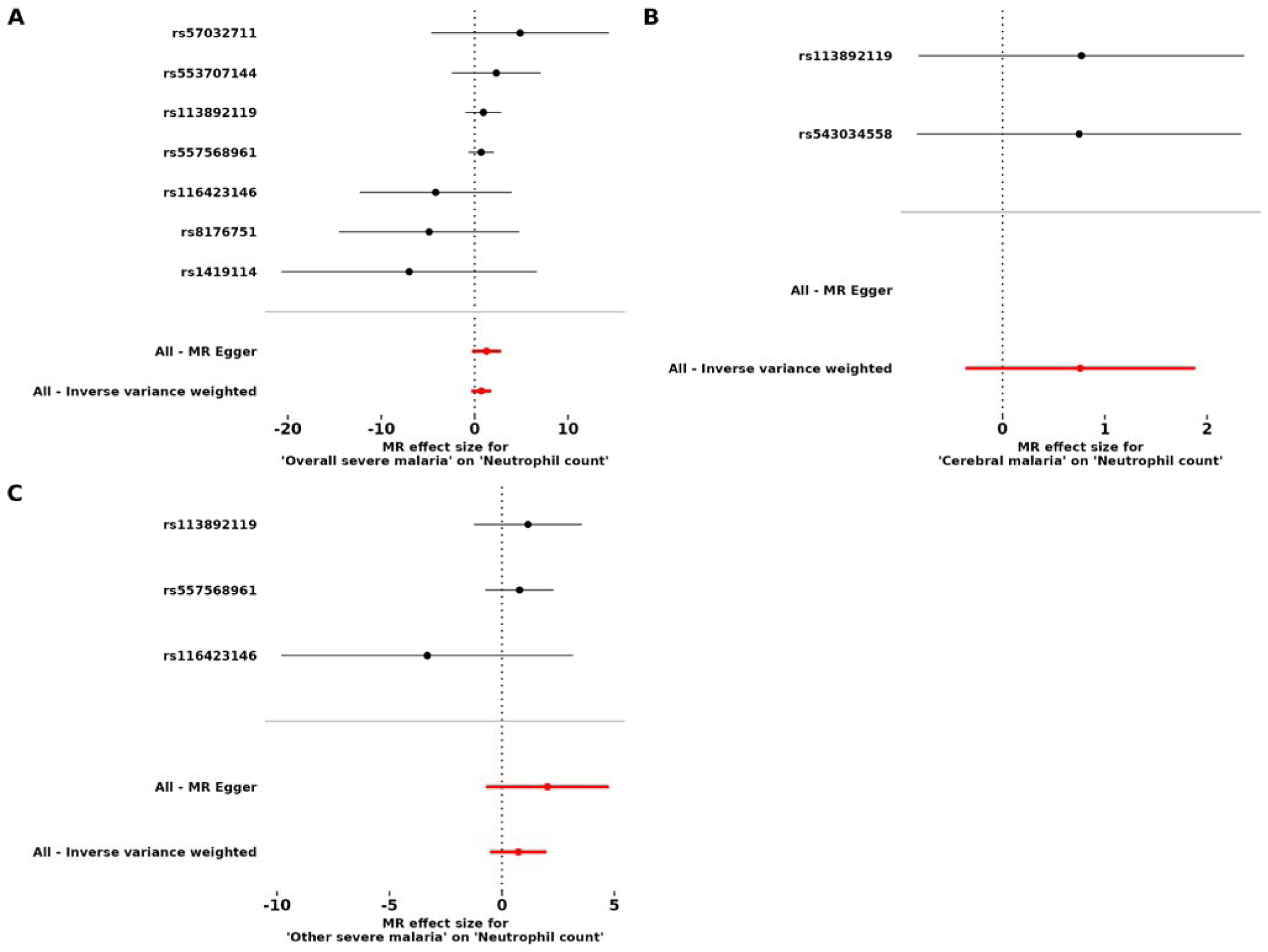
Single-SNP MR analysis of severe malaria on neutrophil count. SNPs proxying for liability to severe malaria are shown on the x-axis. The effect of each SNP proxying for liability to severe malaria is displayed on the y-axis, along with 95% CIs. The MR-Egger and IVW MR methods are shown below the single-SNP analysis.

