## Supplementary material for "Investigating a causal role for neutrophil count on *P. falciparum* severe malaria: a Mendelian Randomization study": Suppementary Methods

SUPPLEMENTARY METHODS

**Pre-GWAS investigative analyses**

Descriptive analyses of nc_log were performed. To study the amount of potential population admixture in the AFR_CAG that could affect the test statistics from a GWAS, an analysis was conducted in R on the Duffy SNP rs2814778 [1] for each Kpop. The preponderance of the Duffy SNP rs2814778 allele distribution was outlined in a PCA plot (PC1~PC2), and its association with nc_log in the AFR_CAG dataset was studied with and without PCs. Tracy-Widom statistics [2] were computed to estimate the number of PCs that are significant i.e. that could be added into the GWAS as covariates. Finally, a power calculation was done assuming a linear model GWAS on the AFR_CAG sample to discuss if there would be enough power to detect a signal.

**SNPTEST and META GWAS**

To test whether the effect estimates from the BOLT-LMM GWAS were biased due to residual population structure that would characterise a population from the African CAG, a number of “sensitivity” GWAS were conducted. This was done with SNPTEST using a linear model algorithm [3,4], with 16 GWAS conducted as follows: 8 GWAS were run on each K-means cluster (Kpop) + the whole sample with the same parameters as in the BOLT-LMM run, and another 8 in the same manner, but with rs2814778 as an additional covariate. To minimise the chance of errors and to reduce the time needed to run each GWAS, a linear model was first conducted in R using the command “lm(nc_log ~ my_covariates)”. The residuals were then pulled with the “residuals()” function and 16 GWAS were run on AFR_CAG with SNPTEST. The Kpop GWAS were then meta-analysed with META [5] under an inverse-variance method based on a fixed-effects model. The result were two meta-analyses: one without accounting for the Duffy SNP rs2814778 called “META-WOD”, and one where the Duffy SNP was included as a covariate, called “META-WD”.

**Conditional & joint association analysis**

GCTA-COJO [6,7] was employed to identify independent signals from the BOLT-LMM GWAS, as well as detect any possible secondary signals arising from a stepwise selection model. SNPs which are close together are usually in LD i.e. their alleles are not random, but correlated [8]. Before running GCTA-COJO, genetic variants with an INFO score < 0.3 were filtered out of the AFR_CAG dataset with QCTOOL. PLINK was then used on this resulting output to filter out related individuals. Following this step, GCTA-COJO was run on the AFR_CAG filtered dataset to identify causal SNPs. These were referred to as “index” in the text. Plots similar to those generated by LocusZoom [9,10] were created in R with the “LocusZooms” package [11].

**Genomic inflation**

The genomic inflation factor lambda (λ) [12] was calculated for the BOLT-LMM and SNPTEST meta-analysis runs. This was complemented by generating quantile-quantile (QQ) [13] to investigate any early deviation of the expected P-values from the observed. Additionally, a Manhattan plot [13] was generated to highlight the BOLT-LMM index SNPs, and two more plots were created to mirror the BOLT-LMM signals with those from a GWAS of neutrophil count in people of African [14] and European [15] ancestry.

**Characterization of functional loci**

A query was placed through the variant effect predictor (VEP) [16] and FUMA [17] on the SNPs in the AFR_CAG filtered dataset. A further, broader literature search was conducted on the index and MR clumping SNPs using Ensembl [18], GeneCards [19], GWAS Catalog [20], The Human Protein Atlas [21], and the Genotype-Tissue Expression (GTEx) project [22].

**Heritability analysis**

An analysis was conducted with GCTA to estimate the proportion of variance in neutrophil count explained by all genetic variants present in the filtered AFR_CAG dataset [23]. First, a power calculation was done to assess whether the sample-size of unrelated people with neutrophil count data (N=5509) would be enough to detect genetic covariance [24]. Default power calculation parameters were used: α = 0.05, *h*^2^ = 0.3, var π = 2e-5; α = P-value significance threshold, *h*^2^ = combined genetic heritability for the trait, var π = variance of the off-diagonal elements of a genetic relationship matrix (GRM) [24]. Afterwards, a GRM was generated from the whole filtered AFR_CAG with the following command. A GRM is essentially a matrix with *n* rows and *y* columns, where n = number of individuals in sample and y = number of SNPs [25]. The *ny* matrix contains the minor allele counts for each SNPs of each individual [25], and it is used by LMM GWA software to adjust for population relatedness that can bias traditional linear model GWA analyses [26]. UKBB phenotypic data was then used to run GCTA-GREML, with and without adjusting for the Duffy SNP rs2814778. Yang et al. propose a way for estimating heritability while accounting for potential LD bias [27]. In brief, segment-based LD scoring was done on each chromosome. SNPs were stratified in R by LD scores in four groups for each chromosome [28], yielding 88 SNP groups in total. A GRM was generated for each SNP group, and GCTA-GREML was run similarly to the previous run.

**GWAS with additional covariates**

Several analyses were conducted to investigate and describe the phenotypic data in the AFR_CAG dataset. Descriptive statistics for neutrophil count were generated to provide information on the sample that the GWAS were run on. Missing data for additional variables were investigated, and an analysis was conducted to test whether missing data in each of these variables showed evidence of affecting neutrophil count. Moreover, a univariable, multivariable ANOVA type II and multivariable ANOVA type III were conducted to assess the variance explained by environmental, multifactorial and immutable (e.g. place of birth) variables. Following the results from the descriptive analyses, another GWAS was run in BOLT-LMM. “Genetic sex”, “time since last menstruation” and “menopause” variables were combined in a single discrete variable called “menstrual_status” and was created as follows: males, quartiles 1-4 of days since last menstruation, menopause, had hysterectomy. The covariates used in this run were sampling device, sample year, sample month, sample day, minutes passed in sample day, UN region of birth, K-means cluster, smoking status, alcohol drinker status, “menstrual_status”, age, body mass index and PCs 1 to 100. 669 individuals were filtered out for missing values and/or preferred not to answer in these variables, bringing the sample-size to 5,310.

**Description of working environment**

All analyses were performed in a Linux environment supported by the University of Bristol’s Advanced Computing Research Centre (ACRC) using the following publicly available software packages: PLINK v1.9 and v2.0 [29,30], QCTOOL v2.0.7 (https://www.well.ox.ac.uk/~gav/qctool/), LDSC v1.0.1 [31], SNPTEST v2.5.4 [3,4], BOLT-LMM v2.3.6 [32], META v1.7 [5], METAL v2011-03-25 [33–35], and GCTA v1.94.0 [6]. All other scripts, analyses, and figures were run and generated in the R environment using version 4.1.2 (Bird Hippie) [36] and Python environment using version 3.7.7 [37] on the ACRC computer clusters.

37	The Python Language Reference — Python 3.7.13 documentation.
